# The Role of Interoception in Lifestyle Factors: A Systematic Review

**DOI:** 10.1101/2024.10.02.24314756

**Authors:** Jesper Mulder, Mirte Boelens, Laura A. van der Velde, Michelle Brust, Jessica C. Kiefte-de Jong

## Abstract

**Background:** Interoception, the perception of the internal state of the body, offers an underrepresented and intriguing potential avenue for altering lifestyle-related behaviors. Interoception is intrinsically related to maintaining homeostasis and the flexible allostatic regulation of complex demands. Internal bodily states may also be influenced by lifestyle factors and related problems. This systematic review aimed to provide insight into the current state of evidence about the role of interoception in lifestyle factors.

**Methods:** Studies from three databases (PubMed, Embase, and Web of Science) were screened for eligibility based on two inclusion criteria: 1) at least one measure of interoception (e.g., interoceptive accuracy, attention, or awareness), and 2) at least one measure of a lifestyle factor (i.e., alcohol consumption, cognitive leisure and relaxation activities, eating behavior, exercise, sleep, or smoking). Risk of bias was assessed using an adjusted quality score checklist, consisting of five items related to study design, population size, exposure measurement, outcome measurement, and adjustment for confounders.

**Results:** The review included 73 studies. Out of the included studies, 51 measured interoceptive accuracy and 36 measured interoceptive attention. Six studies quantified interoceptive awareness. In terms of lifestyle factors, 26 studies on cognitive leisure and relaxation activities, 19 studies on eating behavior, 12 studies focused on alcohol consumption, 10 studies on exercise, five studies on smoking, and four studies on sleep were identified. Findings show positive relations between interoception and cognitive leisure and relaxation activities, eating behavior, and exercise.

Conversely, the consumption of alcohol is negatively related to interoception. Studies in the smoking and sleep lifestyle factors were sparse and of varying methodological quality.

**Discussion:** The findings of this review show that interoceptive domains are associated with various lifestyle factors, but the quality of the evidence is limited. Future longitudinal studies with more objective measures of interoception are needed in order to better understand the interrelations between interoception and lifestyle factors.

**Highlights:**

- Strong support for importance of mindfulness in interoception
- Less strong positive relations between interoception, eating behavior, and exercise
- Less strong negative relation between interoception and alcohol consumption
- Results show lack of articles using objective interoceptive attention measures
- Results show lack of quantification of interoceptive awareness

## 1. Introduction

According to the World Health Organization (WHO), noncommunicable diseases (NCDs), including cardiovascular diseases, cancers, diabetes, and chronic respiratory diseases, account for 74% of global deaths. These NCDs are directly influenced by lifestyle factors [1]. Lifestyle factors can be defined as a set of modifiable behaviors and habits of individuals in their daily lives. As the WHO aims to reduce premature deaths from NCDs by one third before 2030, exploring new directions for prevention and treatment is crucial for figuring out the most effective strategies to support lifestyle changes. The adaptation of lifestyle behaviors can be challenging for many individuals, since many biosocial factors play a role in behavioral change [2]. Biosocial factors are dynamic and bidirectional biological and sociological aspects, relationships, and contexts, which comprise the processes of human development. One of these aspects is interoception, which offers an underrepresented and intriguing potential avenue for altering lifestyle-related behaviors.

Interoception, the perception of the internal state of the body (e.g., heartrate, sweating, or hunger), is a field of research that has received increasing scientific interest over the years [3]. Research into interoception has demonstrated the importance of internal bodily signals for physical and mental well-being, as processing these signals helps maintain homeostasis (i.e., maintenance of physiological stability) [4]. Additionally, interoception is intrinsically related to the predictive control of bodily signals, both in maintaining physiological balance and in the flexible allostatic regulation of complex demands [5]. For example, when the immune system communicates states of infection and inflammation to the brain, this happens via interoceptive pathways [6]. The body responds to these states by activating cardiovascular and gastrointestinal reflexes, regulating peripheral immune reactions, and patterns of responses called sickness behaviors (i.e., fatigue, reduced food and water intake, social isolation, and fever) [7–9]. This illustrates how changes in internal bodily states could affect behaviors, mediated by interoceptive pathways.

These internal bodily states may also be influenced by lifestyle factors and related problems. An example is the role of interoception in exercise. If there is a mismatch between perceived fatigue communicated through interoceptive pathways, and the actual physiological state, this can result in hypoactivity (i.e., reduced participation in exercise, which may lead to problems related to sedentary behavior) or hyperactivity (i.e., continuation of physical exercise to the point of reaching dangerous physiological boundaries) [10]. Another example is the role of interoception in eating behavior.

During eating, changes in hunger and satiety signals determine eating responses and maintain homeostasis. When hunger and satiety signals are not adequately perceived or acted on, this may lead to overeating and lifestyle-related diseases such as obesity [4]. Lifestyle-related diseases and disorders shape variations in the response of the immune system [11], and a healthy lifestyle is regarded as an important factor in strengthening the immune system in the prevention and treatment of diseases [12]. When an individual’s physical and mental well-being is affected by lifestyle-related problems, similar reactions as with infection and inflammation may occur [7–9]. Research into the underlying mechanisms of interoception has yielded valuable insights in relation to lifestyle factors. Previous work concentrated mainly on the role of interoception in various conditions, such as alexithymia, sickness behaviors, fatigue, depression, eating disorders, autism spectrum disorders, and anxiety [5, 13, 14]. However, to the best of our knowledge, the relationship between interoception and lifestyle factors has not yet been systematically evaluated, particularly with regard to the quality of the studies involved.

We aim to provide insight into the current state of evidence about the role of interoception in lifestyle factors by conducting a systematic review. The overall goal is to synthesize findings and identify general patterns in the relation between interoception and lifestyle factors. Insights into the association between interoception and a healthy lifestyle can be used to promote a healthy lifestyle through the development and improvement of lifestyle interventions focusing on interoception.

## 2. Methods

### 2.1. Eligibility criteria and search strategy

A protocol for this systematic review was developed in line with the Preferred Reporting Items for Systematic Reviews and Meta-Analyses (PRISMA) for protocols guidelines, and registered prospectively within PROSPERO with registration number CRD42023465957. This systematic review of literature was performed according to the PRISMA statement [15]. The review included peer-reviewed papers written in English, with no limitations on publication date. Only quantitative studies were included. Evidence syntheses (e.g., reviews) or non-original research articles (e.g., commentaries) were excluded. This review included adults as population of interest (i.e., ≥ 18 years of age). However, if results for both adults and children were presented separately, the study was included and only the results for the group of adults were extracted. Studies were screened for eligibility based on two inclusion criteria: 1) the study had to include at least one measure of interoception (e.g., interoceptive accuracy, attention, or awareness), and 2) at least one measure of a lifestyle factor (i.e., alcohol consumption, cognitive leisure and relaxation activities, eating behavior, exercise, sleep, or smoking). Studies were excluded when 1) the study focused on alexithymia, emotion, or autism spectrum disorders, 2) eating disorders such as anorexia nervosa, bulimia nervosa, binge eating disorder, and emotional eating (a recent review already covered the relation between interoception and eating disorders) [16], and 3) when the population consisted exclusively of individuals with impaired cognitive function, hypertension, obesity, or other non-lifestyle related disorders.

A systematic literature search was conducted from the date of inception to the 21^st^ of September 2023 (date of last search) in three databases (PubMed, Embase, and Web of Science). The search strategy was developed using medical subheadings and free-text words in the title or abstract and enhanced by a medical librarian with expertise in systematic reviews. Search strategies were adapted to fit each database. The search strategy included terms such as interoception (accuracy, attention, awareness, sensitivity, or sensibility), as well as lifestyle factors (alcohol consumption, cognitive leisure and relaxation activities, eating behavior, exercise, sleep, or smoking). The complete search string can be found in Appendix A. Additionally, reference lists of included articles were screened by hand.

### 2.2. Selection and data extraction

Records were imported into EndNote [17] to check for duplicates. Titles and abstracts were independently screened for eligibility by at least two authors, and by using the automation tool ASReview [18]. ASReview uses an active researcher-in-the-loop machine learning algorithm to rank articles from high to low relevancy for inclusion by text mining. The algorithm is able to detect 95% of the final included studies within the first 20% of studies that are presented to the reviewer, significantly reducing the time spent screening titles and abstracts while maintaining quality [19]. To provide relevant terms to the automation tool, a few systematic reviews and meta-analyses were entered at the beginning of the screening process. These review articles were not included in the eligibility assessment phase. Data were independently extracted by at least two authors, using a manually created data extraction form. Extracted information included study characteristics (e.g., title, first author, year, and study design), participant demographics (e.g., number of participants, sex, ethnicity, country, setting, and other population characteristics), outcome measures of interoception and lifestyle factors (e.g., interoception domain and measure according to the 2×2 factorial structure for interoception shown in Figure 1, and lifestyle factor and measure), results of the measurements (e.g., statistical analyses, confounders, interoception results, lifestyle factor results, and correlations), and main findings reported in the article. In cases of discrepancies between authors, issues were resolved by discussion until consensus was reached or by involving a third author.

**Figure 1.**
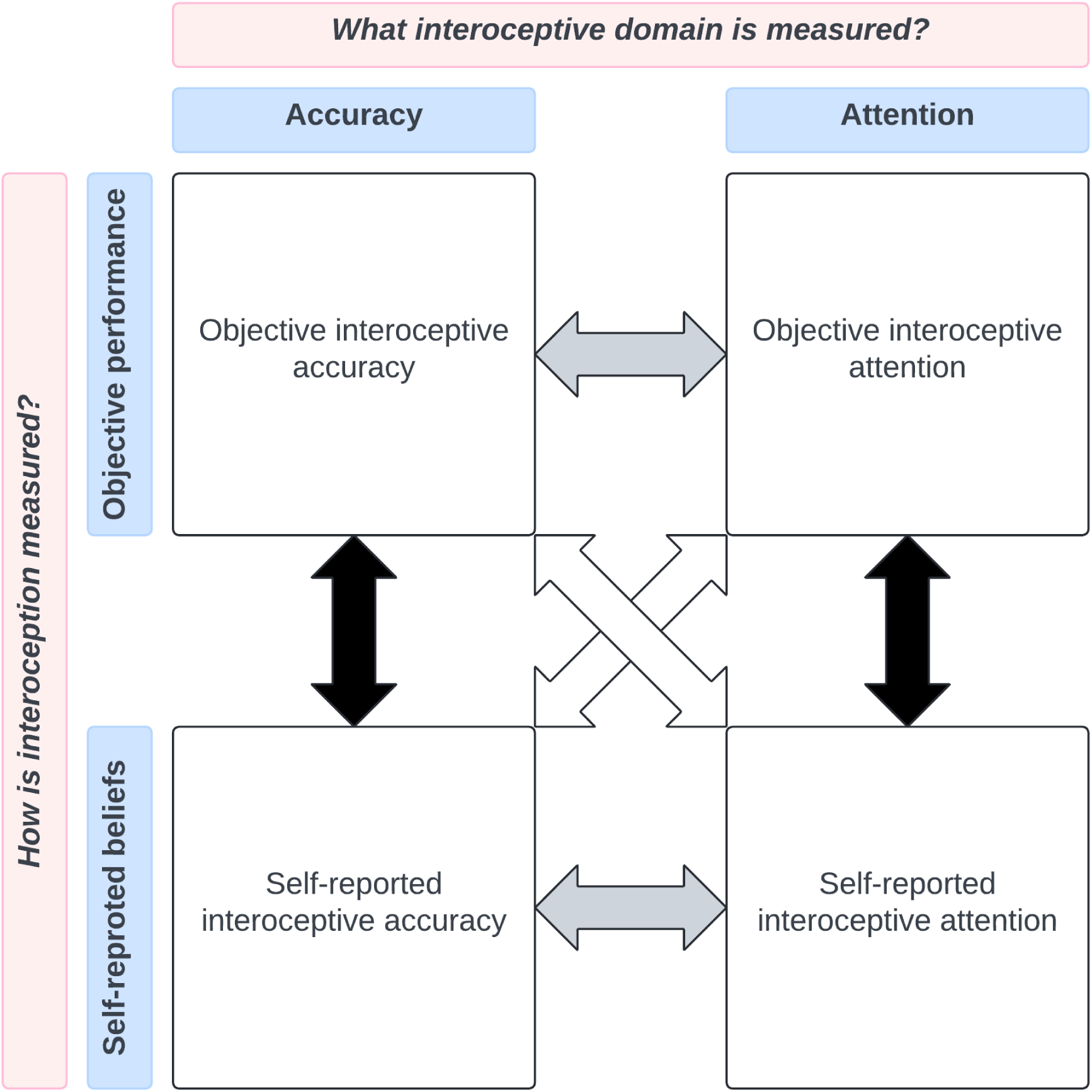
Simplified version of the 2×2 factorial structure for interoception developed by Murphy et al. 2019 [3]. The quadrants represent interoceptive accuracy assessed with objective performance tasks (top left), interoceptive attention measured with objective performance tasks (top right), interoceptive accuracy assessed with self-report measures (bottom left), and interoceptive attention assessed with self-report measures (bottom right). The arrows represent the interaction between objective and self-report measures (i.e., interoceptive awareness; black arrows), interoceptive accuracy and attention (grey arrows), and across different interoceptive domains and measures (white arrows)

### 2.3. Synthesis methods

To categorize interoceptive domains and measures, the 2×2 factorial structure for interoception developed by Murphy and colleagues was adopted (Figure 1) [3]. The first factor in this model makes the distinction between interoceptive accuracy (i.e., an individual’s ability to perceive interoceptive signals accurately) and attention (i.e., the degree to which interoceptive signals are objects of attention). The second factor distinguishes between the way interoceptive accuracy or attention is measured: an objective performance measure or a self-report measure. Subsequently, interoceptive awareness can be quantified by assessing the correlation between self-reported beliefs and objective measures.

We operationalized lifestyle factors based on the following definitions: 1) alcohol consumption, which included all behavior related to the consumption of alcoholic beverages [20]; 2) cognitive leisure and relaxation activities, which included leisure activities such as reading, writing, playing games like cards, checkers, crosswords or other puzzles, participating in groups or clubs, attending movies, plays, lectures or public meetings, or playing music, and stress or anxiety reducing activities such as mindfulness, meditation, yoga, and tai-chi [21–24]; 3) eating behavior, which included food choice and motives, feeding practices, and problems related to eating [25]; 4) exercise was defined as regular physical activity with the intention of improving physical fitness or health, which ranges from low intensity exercise a few times a week to participating in daily high intensity exercise [26]; 5) sleep included aspects related to sleep such as sleep duration, quality, latency, and stages [27]; and finally, 6) smoking behavior, which is the action or habit of inhaling and exhaling smoke from burning substances [28]. For each of these lifestyle factors, the 2×2 factorial structure for interoception was used to create harvest plots.

### 2.4. Study risk of bias and quality of evidence assessment

Risk of bias was assessed using an adjusted quality score checklist, based on three other checklists, to align with the objectives of the current systematic review and to accommodate both interventional and observational study designs [29–31]. The quality score consists of five items related to study design, population size, exposure measurement, outcome measurement, and adjustment for confounders (see Appendix B). Each item was scored 0, 1, or 2, allowing for a total score between 0 and 10, with 10 representing the highest quality [29–31]. Quality scores were assigned to all included articles by at least two authors. In case of discrepancies in quality score assessments between the two authors, the studies were discussed until consensus was reached, or a third author was consulted. Studies were considered of low, moderate, or high methodological quality when the quality score was ≤ 3, 4 – 6, or ≥ 7, respectively. Information on the quality and findings of the studies were combined to rate the level of evidence, based on the rating system shown in Table 1. The level of evidence was categorized as strong (i.e., three studies of high OR five studies of moderate quality with/without statistically significant effects), moderate (i.e., two studies of high OR three studies of moderate quality with/without statistically significant effects), limited (i.e., one study of high OR two studies of moderate quality with/without statistically significant effects), or conflicting (i.e., < 2:1 ratio between studies with and without statistically significant effects).

**Table 1.** Level of evidence ratings.

| Level of evidence | Definition |
| --- | --- |
| Conflicting | Conflicting results (< 2:1 ratio) between studies finding (no) discriminatory, explanatory and/or predictive effects |
| Limited | One study of high OR two studies of moderate quality finding (no) discriminatory, explanatory and/or predictive effects |
| Moderate | Two studies of high OR three studies of moderate quality finding (no) discriminatory, explanatory and/or predictive effects |
| Strong | At least three studies of high OR at least five studies of moderate quality finding (no) discriminatory, explanatory and/or predictive effects |

## 3. Results

A total of 4,609 studies were identified. After removing duplicate records (n = 1,603) and excluding studies based on title and abstract (n = 2,564), 442 studies remained for full-text screening. Five additional studies were identified through reference list screening. Assessment of full-text studies and identification of studies via other methods resulted in 73 studies included in the review (see Figure 2 for the selection process).

**Figure 2.**
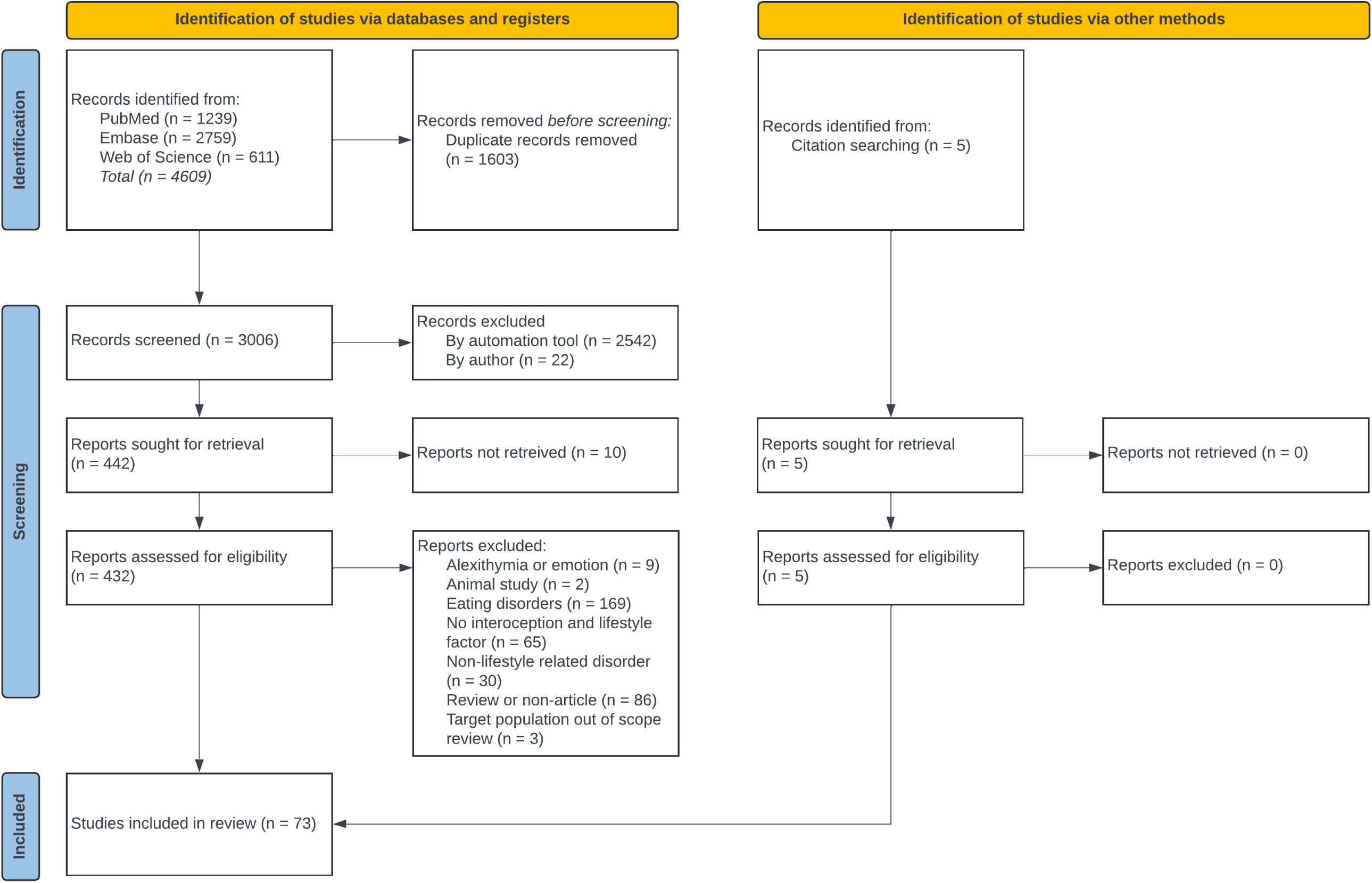
Flow-chart showing the study selection process.

Table 2 provides an overview of the study characteristics of the included studies. The included studies involved 10,695 participants, ranging from seven to 1,447 participants per study. Our review covered a total of 33 observational studies [32–64], 20 randomized controlled trials [65–84], and 20 quasi-experimental studies [85–104].

**Table 2.**
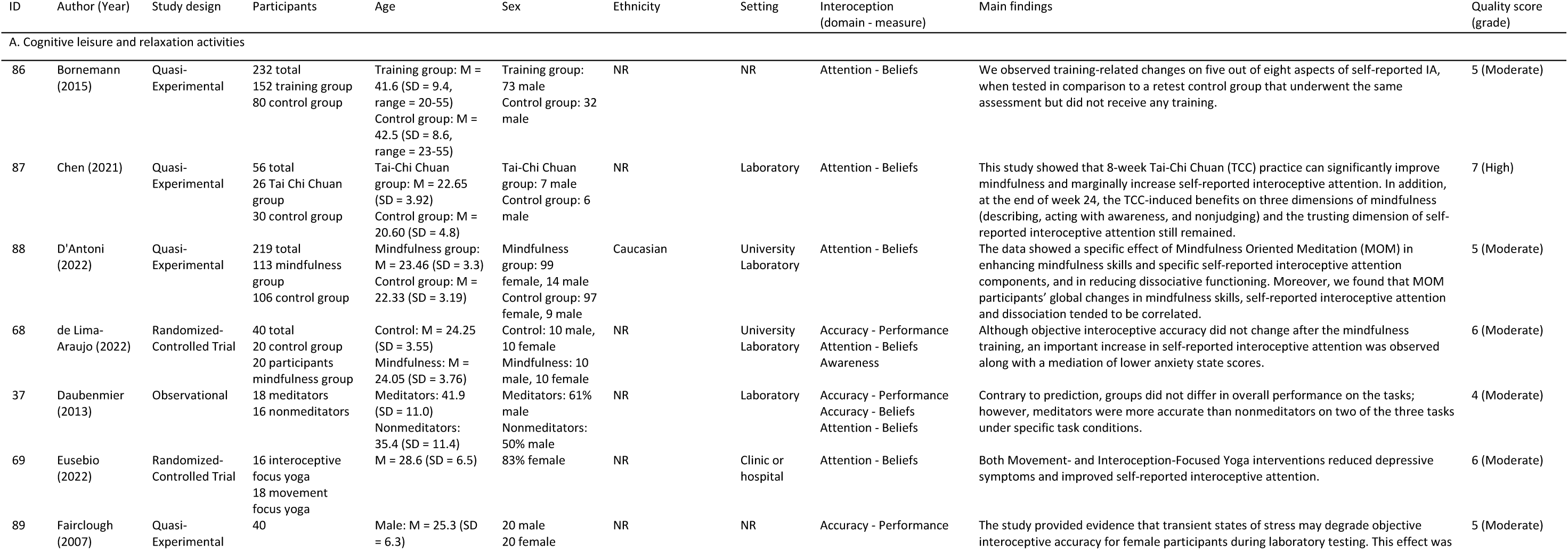
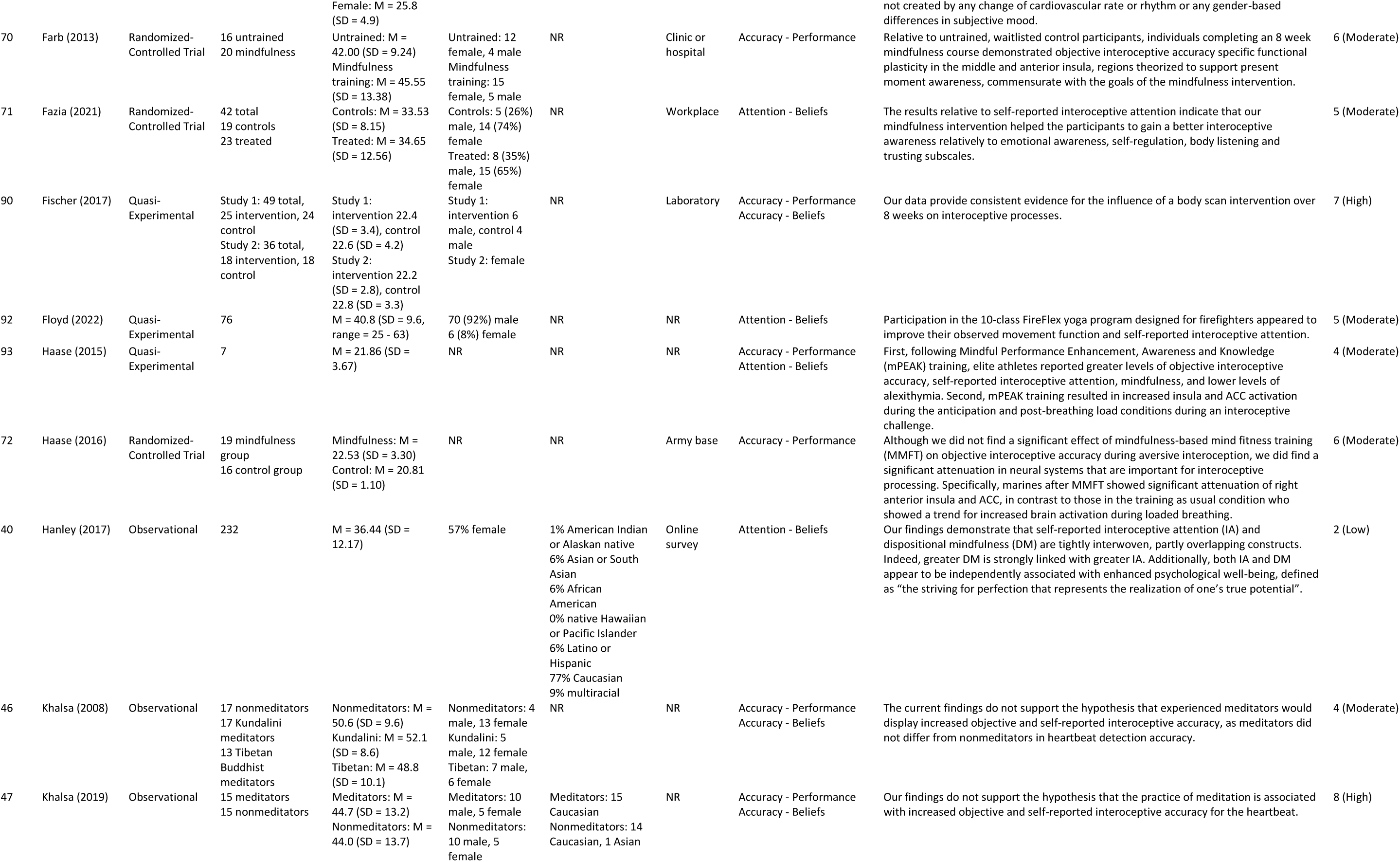
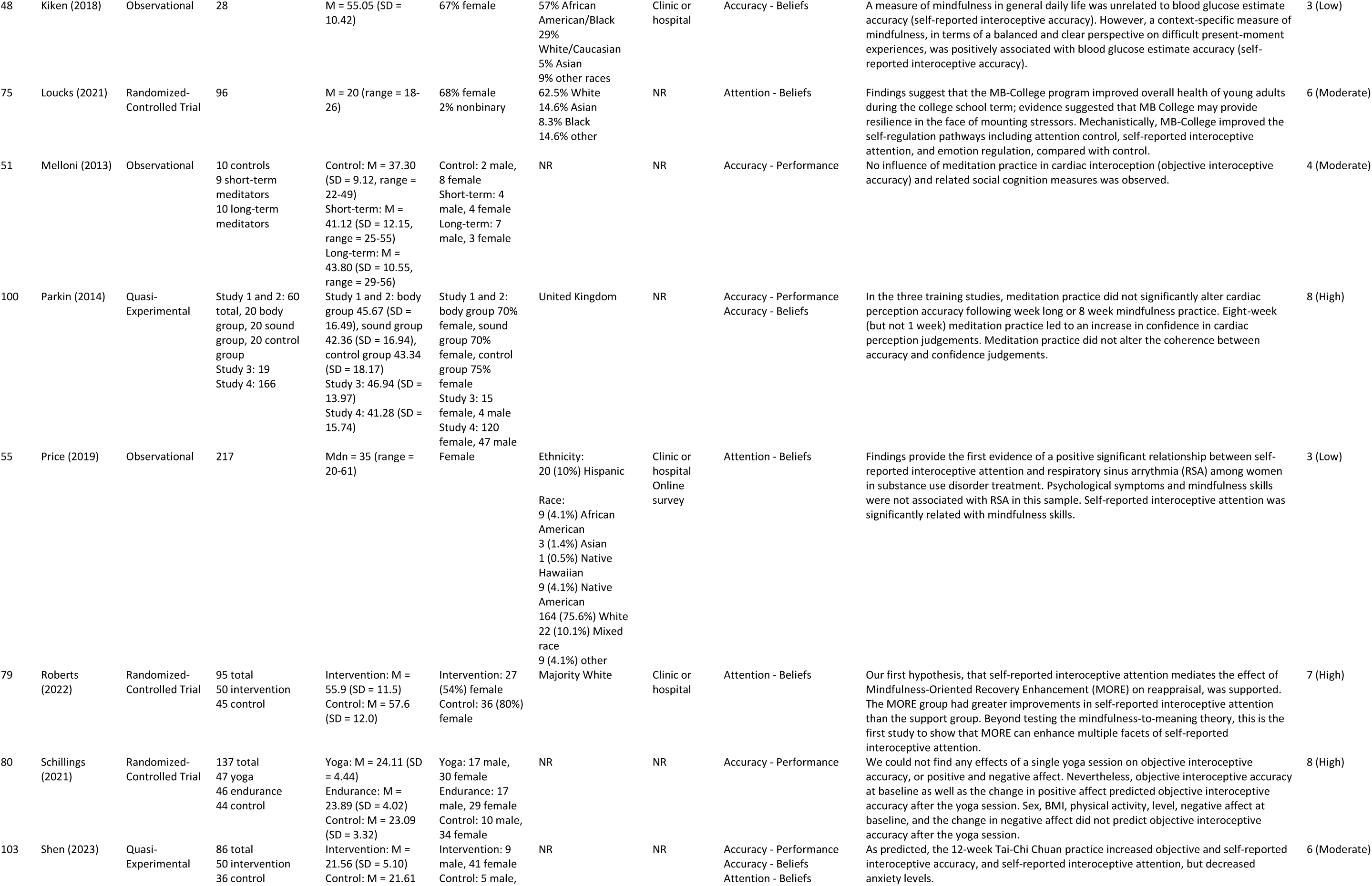
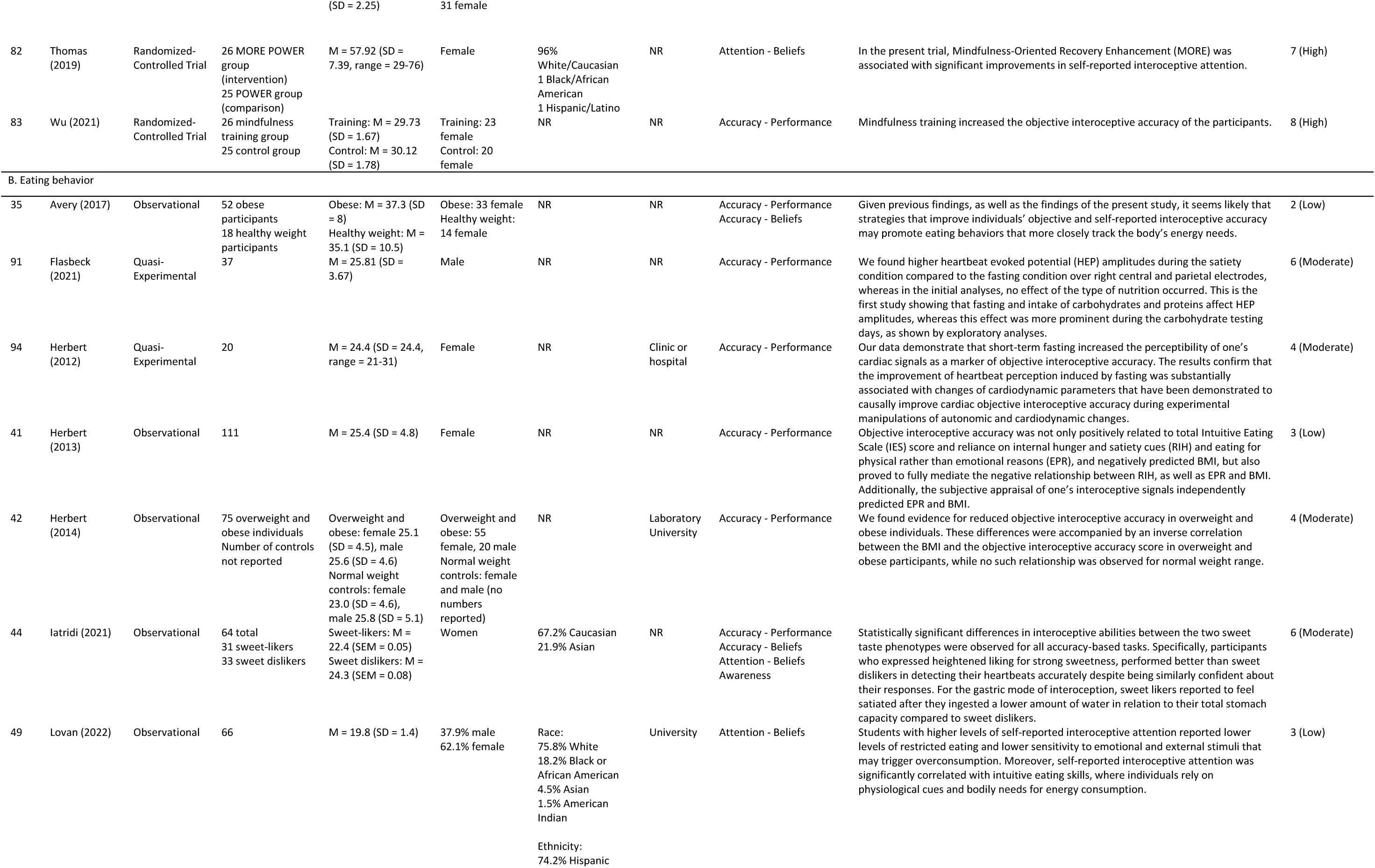
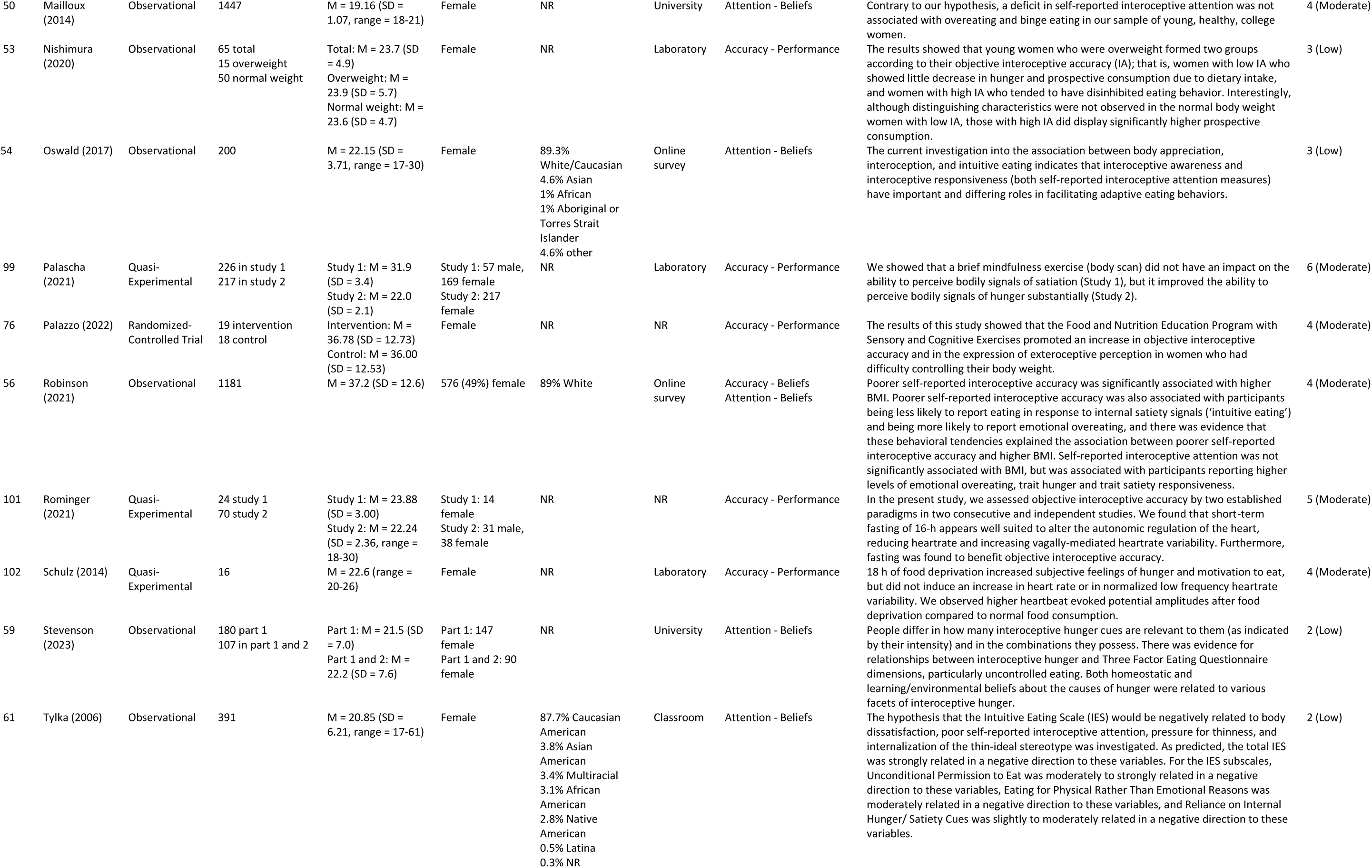
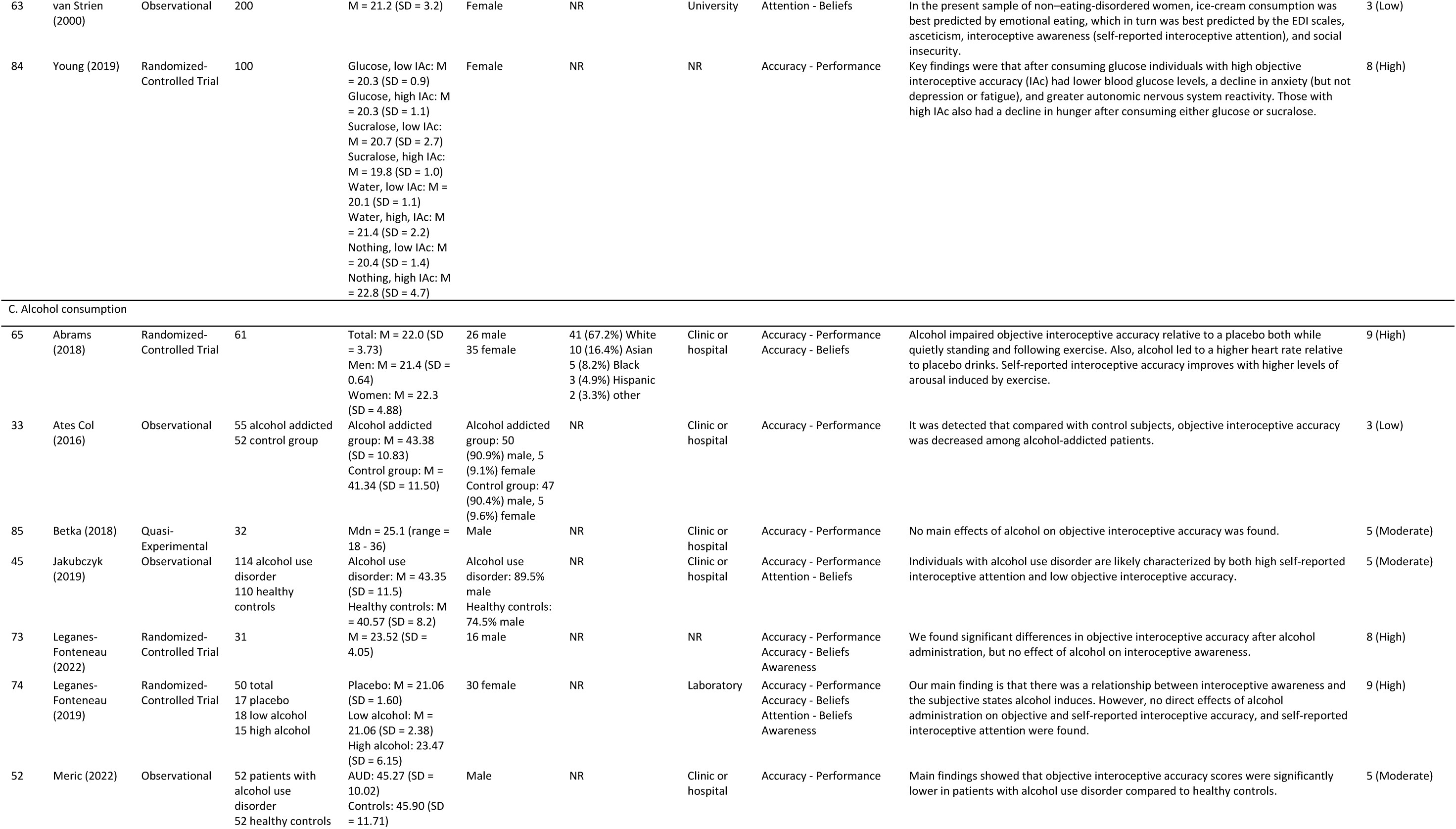
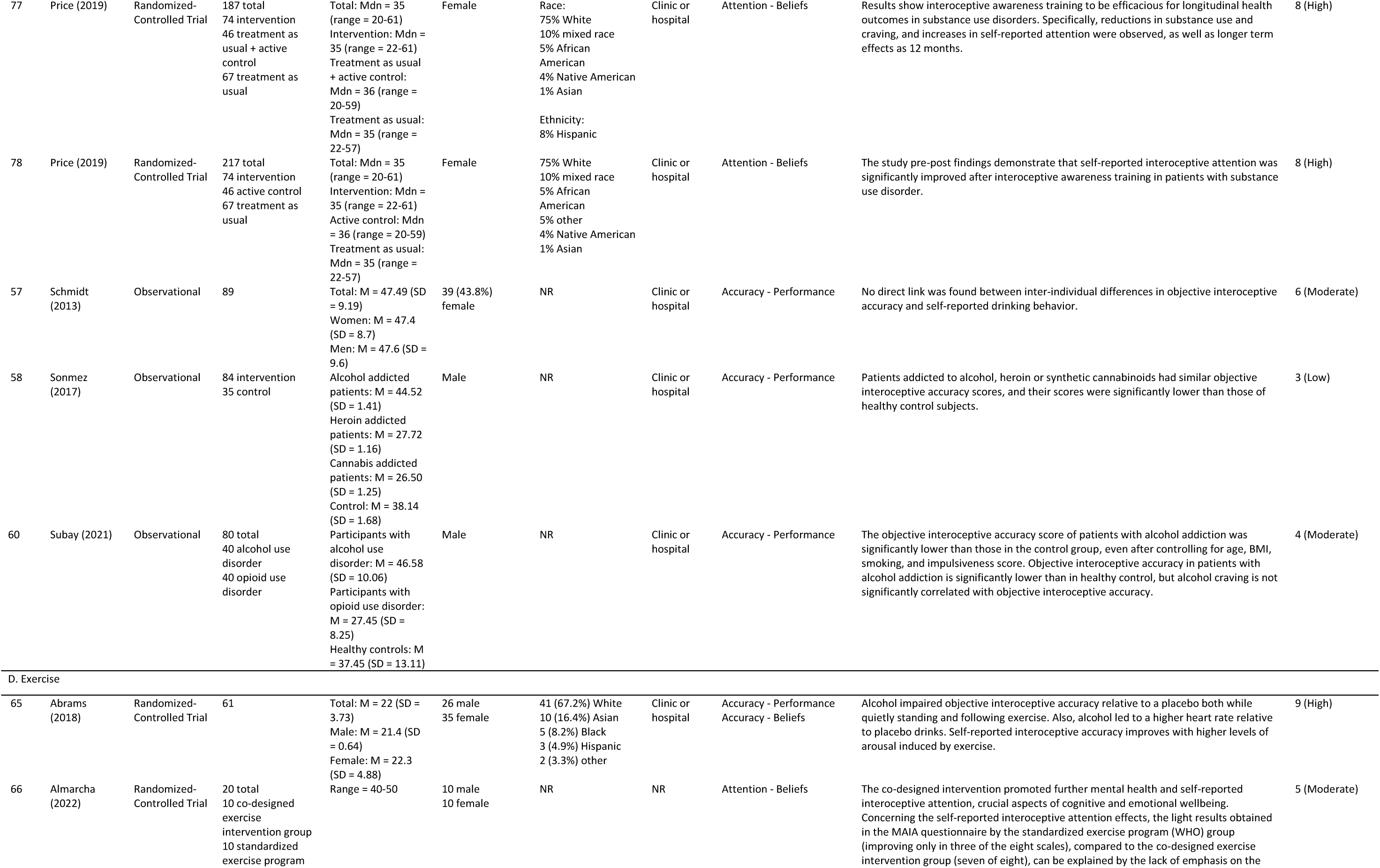
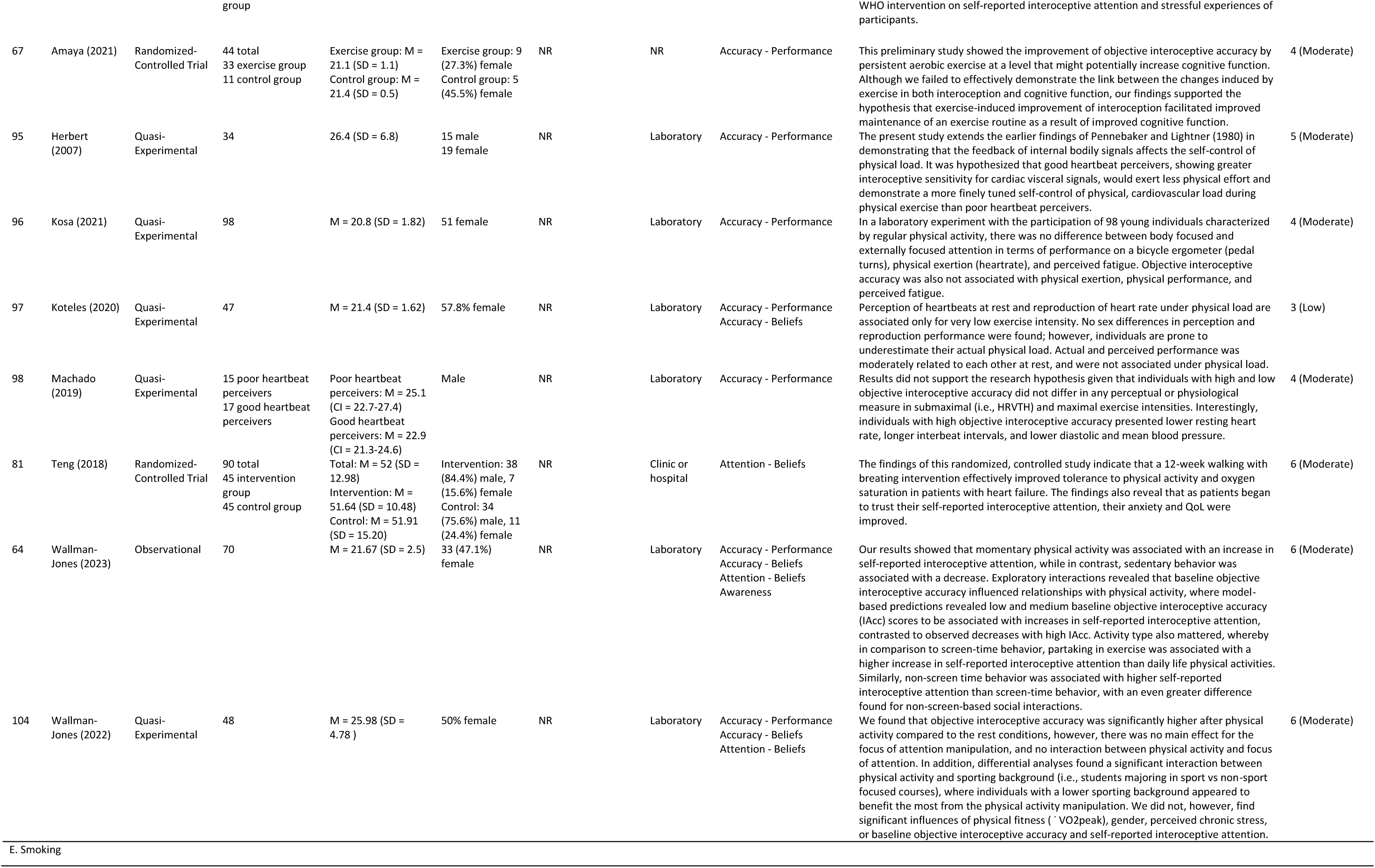
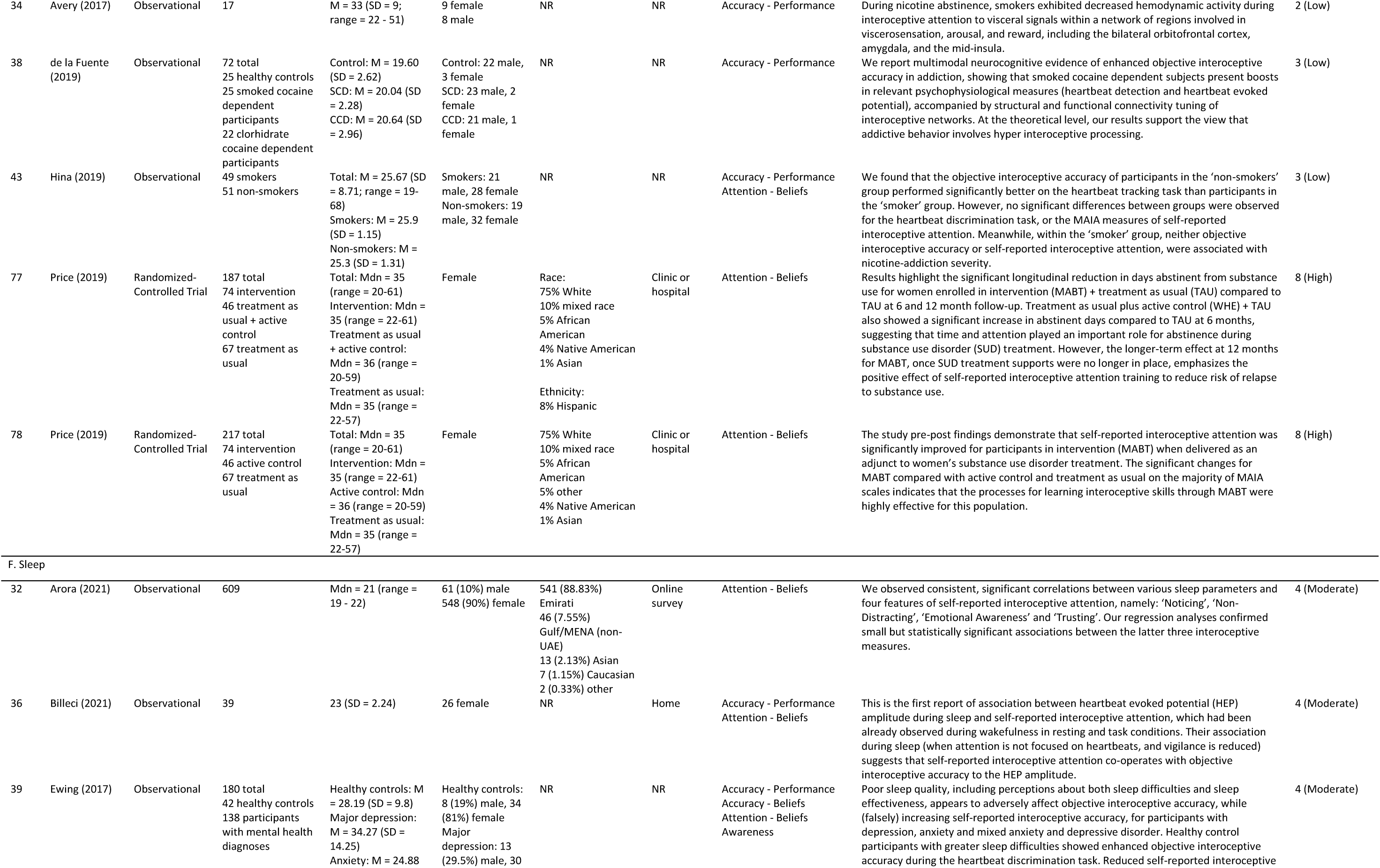
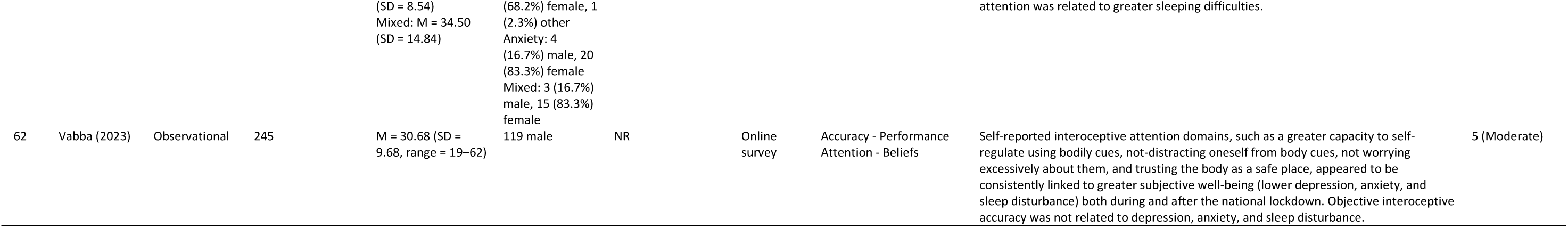
Summary table of included studies per lifestyle factor (A-F), including reference ID, first author (year of publication), study design, number, age, sex, and ethnicity of participants, study setting, interoceptive domain and measure, main findings, and study quality score (grade). M = Mean, SD = Standard Deviation, Mdn = Median, NR = Not Reported.

Harvest plots to visualize the distribution of included studies over interoceptive domains and lifestyle factors are shown in Figure 3. Out of the included studies, 51 measured interoceptive accuracy, with 49 instances of objective performance measures [33–39, 41–47, 51–53, 57, 58, 60, 62, 64, 65, 67, 68, 70, 72–74, 76, 80, 83–85, 89–91, 93–104] and 17 self-report measures [35, 37, 39, 44, 46–48, 56, 64, 65, 73, 74, 90, 97, 100, 103, 104]. Interoceptive attention was assessed in 36 studies, all of which used self-report measures [32, 36, 37, 39, 40, 43–45, 49, 50, 54–56, 59, 61–64, 66, 68, 69, 71, 74, 75, 77–79, 81, 82, 86–88, 92, 93, 103, 104]. Six studies quantified interoceptive awareness (i.e., the interaction between interoceptive accuracy and attention) [39, 44, 64, 68, 73, 74]. In terms of lifestyle factors, 26 studies on cognitive leisure and relaxation activities, 19 studies on eating behavior, 12 studies focused on alcohol consumption, 10 studies on exercise, five studies on smoking, and four studies on sleep were identified. Some studies included more than one interoceptive domain, interoceptive measure, or lifestyle factor and were therefore analyzed in multiple categories.

**Figure 3.**
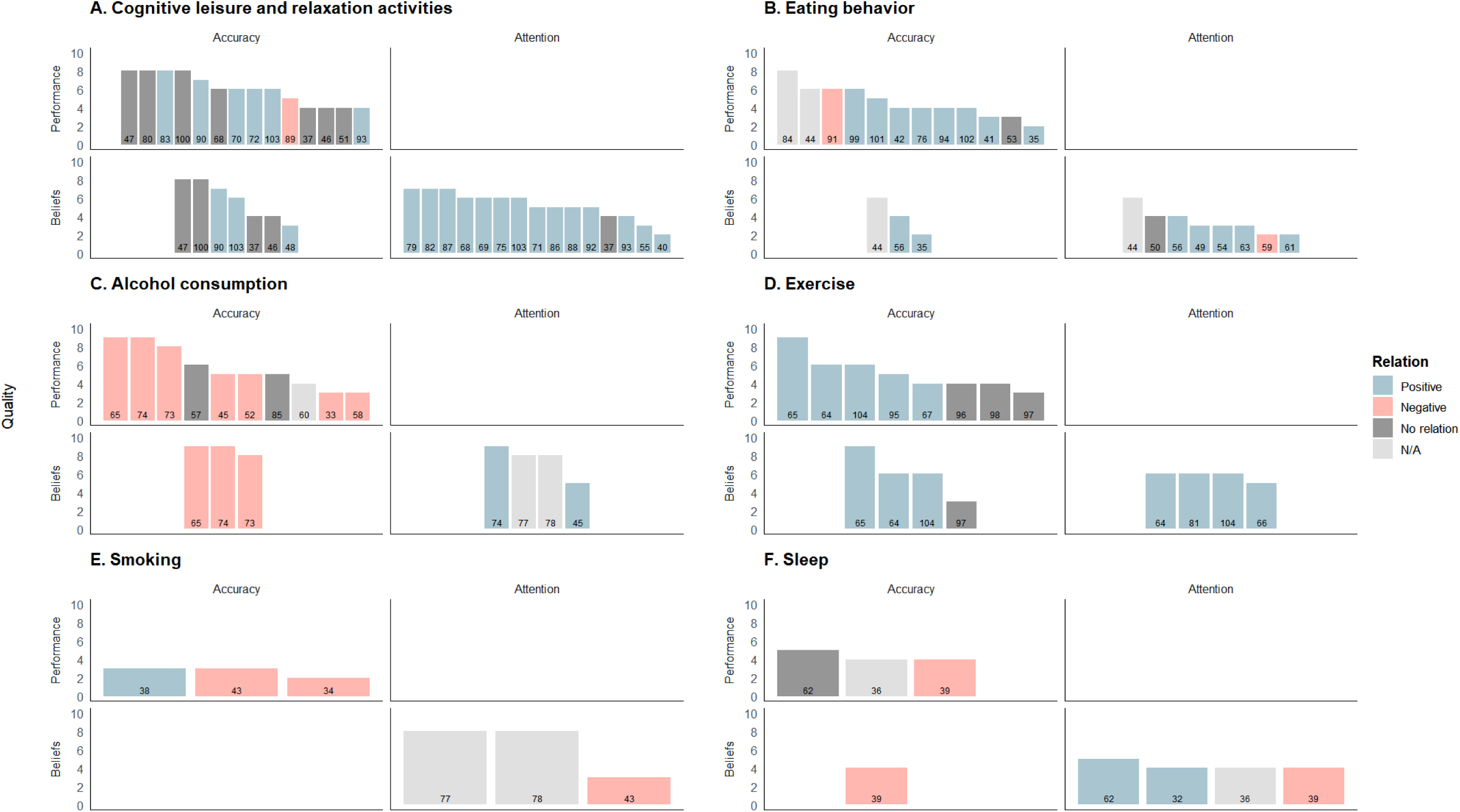
Harvest plots per lifestyle factor. (A-F), based on the 2×2 factorial model for interoception. Each bar represents a single study, with reference numbers within the bar. The height of each bar represents the assigned quality score (0-10), and the color of each bar the direction of the relation between the lifestyle factor and interoceptive domain. Note: some studies measured both interoceptive accuracy and attention, and/or multiple lifestyle factors (e.g., eating behavior and exercise). These articles are therefore represented in multiple lifestyle factors, and/or plots.

Quality scores ranged from two to nine, with a mean quality score of 5.0. We identified 14 studies with high, 42 studies with moderate, and 17 studies with low methodological quality.

### 3.1. Cognitive leisure and relaxation activities [37, 40, 46–48, 51, 55, 68–72, 75, 79, 80, 82, 83, 86–90, 92, 93, 100, 103]

The cognitive leisure and relaxation activities identified in this review were mindfulness, Tai-Chi, yoga, and meditation activities. Strong evidence suggests mindfulness skills and training positively relates to self-reported interoceptive attention. Additionally, moderate evidence was found for the positive relation between mindfulness and objective interoceptive accuracy. There was limited evidence on the positive effect of Tai-Chi practices on self-reported interoceptive attention. Several studies indicated there were no effects of meditation techniques or no differences between meditators and non-meditators in both objective and self-reported accuracy, showing moderate and limited levels of evidence, respectively. Conflicting results were found for the relationship between meditation practices and self-reported interoceptive attention. Both movement– and interoception-focused yoga programs were found to positively relate to self-reported interoceptive attention. Quality scores of the studies within this lifestyle factor ranged from two to eight, with an average of 5.6.

### 3.2. Eating behavior [35, 41, 42, 44, 49, 50, 53, 54, 56, 59, 61, 63, 76, 84, 91, 94, 99, 101, 102]

Positive relations between eating behavior (i.e., intuitive eating and eating behaviors that allow for better tracking of energy needs), objective and self-reported interoceptive accuracy, and self-reported interoceptive attention were observed. A moderate level of evidence was found for the association that fasting increases objective interoceptive accuracy. Additionally, two studies reported that mindfulness-based eating interventions have a positive effect on objective interoceptive accuracy. These studies combined had a limited level of evidence. Eating behavior quality scores averaged 4.0, ranging from two to eight.

### 3.3. Alcohol consumption [33, 45, 52, 57, 58, 60, 65, 73, 74, 77, 78, 85]

Moderate evidence was found on the negative relation between alcohol consumption and objective interoceptive accuracy: most of the studies showed that objective interoceptive accuracy is impaired in individuals suffering from alcohol use disorders, or that the use of alcohol decreases objective interoceptive accuracy scores in healthy individuals. Similar evidence was found for the negative effect of alcohol on self-reported interoceptive accuracy.

Additionally, two studies showed that interoceptive training helps in improving self-reported interoceptive attention, abstinence of substance use, and reducing risk of relapse in substance use disorders (e.g., alcohol, smoking, and other substances), providing a moderate level of evidence. The quality score of alcohol consumption studies had a mean of 6.1 and a range of three to nine.

### 3.4. Exercise [64–67, 81, 95–98, 104]

Based on four studies, a moderate level of evidence was found for the positive relation between exercise and self-reported interoceptive attention. Similarly, based on three studies, a moderate level of evidence indicated a positive relation between exercise and self-reported interoceptive accuracy. A moderate level of evidence was found on the positive relation between exercise and objective interoceptive accuracy: five studies showed a positive relation between exercise and objective interoceptive accuracy, however three studies reported there was no relation between the two. Also, one study mentioned that there was no relation between self-reported interoceptive accuracy and exercise. The majority of studies in this section investigated the effect of acute exercise protocols on interoception. The mean quality score for studies on interoception and exercise was 5.2, with a range from three to nine.

### 3.5. Smoking [34, 38, 43, 77, 78]

Results on the relation between smoking behavior, objective interoceptive accuracy, and self-reported interoceptive attention were inconclusive due to low-quality studies and varying findings. In substance use disorders (e.g., smoking, alcohol, and other substances), interoceptive training can improve self-reported interoceptive attention and abstinence of substance use, while reducing risk of relapse. The quality score from studies in this section averaged 4.8, with a range of two to eight.

### 3.6. Sleep [32, 36, 39, 62]

Limited evidence shows that there is a positive relation between sleep quality and self-reported interoceptive attention. Conversely, the relation between sleep and objective interoceptive accuracy was found to be negative. More sleep difficulties were reported to be related to enhanced interoception scores. The mean quality score for the four studies in the interoception and sleep section was 4.3 and ranged from four to five.

## 4. Discussion

### 4.1. General overview

The aim of the current systematic review was to provide insight into the current state of evidence about the role of interoception in lifestyle factors (i.e., alcohol consumption, cognitive leisure and relaxation activities, eating behavior, exercise, sleep, or smoking). Most of the 73 included studies investigated cognitive leisure and relaxation activities or eating behavior, while fewer studies focused on the role of interoception in alcohol consumption, exercise, smoking, or sleep. More articles focused on interoceptive accuracy (i.e., an individual’s ability to perceive interoceptive signals accurately) than interoceptive attention (i.e., the degree to which interoceptive signals are objects of attention). The instances of objective performance measures and self-reported beliefs were equally distributed. However, while several studies measured both objective performance and self-reported beliefs, only six studies quantified interoceptive awareness.

In general, findings in this review show positive relations between interoception and the lifestyle factors cognitive leisure and relaxation activities, eating behavior, and exercise. Conversely, the consumption of alcohol is negatively related to interoception. Studies in the smoking and sleep lifestyle factors were sparse and of varying methodological quality, making it difficult to determine a clear direction of the relationship of these factors with interoception. Studies in the cognitive leisure and relaxation activities category were mostly experimental in design, suggesting a causal relationship with interoception. In other lifestyle factors study designs were either equally distributed between observational and experimental designs or skewed towards observational designs. Notably, no studies were identified that investigated objective interoceptive attention, suggesting that no methods to measure this domain are available, or the available methods are not deemed reliable, valid, or feasible. Most studies were of moderate or high methodological quality. Below we will discuss the findings for each lifestyle factor in more detail.

### 4.2. Cognitive leisure and relaxation activities

Most studies in this review focusing on cognitive leisure and relaxation activities included an element of mindfulness. Mindfulness is a psychological concept with emphasis on present moment awareness, focus on purpose, and non-judgmental thinking [105]. Mindfulness-based interventions encourage individuals to consciously focus on bodily sensations without judgement, and with openness, curiosity, and self-compassion [106]. In the current review, as well as in previous reviews [106, 107], the main finding in terms of mindfulness is that these practices improve interoceptive accuracy and attention. Emotional-motivational states are strengthened through mindfulness training, and mindfulness practice helps in the development of interoceptive brain regions [40, 48, 68, 71, 75, 82, 86, 88, 90]. Mindfulness interventions have been shown to promote functional connectivity, resulting in greater anterior insula activation [70, 72, 79, 93]. Tai-chi and yoga, types of mindfulness practices that combine physical movement with mental focus, show that combining a focus on the mind with physical movement can enhance the regulative ability of interoception and awareness of internal stimuli [69, 80, 87, 103]. Conversely, meditation, which differs from mindfulness training in that it is more focused on mental and emotional calmness, does not seem to be related to interoception. Several studies identified in this review indicate no difference between meditators and nonmeditators in terms of interoceptive accuracy [37, 46, 47, 51, 100]. One of the explanations for these findings can be found in the heartbeat tracking or discrimination tasks used to assess interoceptive accuracy. Training in meditation traditions is commonly focused on breathing, which might not translate to other bodily sensations such as heartrate [46, 47]. These findings are not in line with other narrative and systematic reviews on mediation and interoception, where increases in size and activation of the insular cortex have been observed in meditators but not in nonmeditators [107, 108]. In general, mindfulness and practices related to mindfulness seem beneficial in the development of interoceptive abilities.

### 4.3. Eating behavior

The results of the current review indicate a positive relationship between interoception and intuitive eating. Intuitive eating uses hunger and satiety cues to guide eating behavior, as opposed to emotions [41]. These findings replicate results from other reviews that show the association of interoception with awareness of hunger and satiety cues, which is used to regulate food intake [109, 110]. After eating, there is an increase in sensitivity to visceral-afferent signals, which signals fullness via hormonal and neural pathways [91]. Even when emotional eaters are able to perceive bodily signals as well as intuitive eaters, it seems emotional eaters respond less positively to these signals or they are not used as much in decision-making processes related to eating behavior [54, 56]. For example, in healthy weight individuals hunger is perceived as a physiological desire for energy, while in obese individuals hunger is perceived as the absence of reward [35]. The connection between interoception and eating behavior is in the insular cortex, which plays a vital role in detecting energy needs [111]. Several studies have indicated activation of brain structures that are relevant for interoception during periods of fasting, showing the relationship between hunger and interoception is found in these areas of the brain [94, 101, 102]. Interestingly, one study showed that individuals with lower interoceptive abilities were more likely to overeat, but also that overweight individuals with high interoception scores were more prone to overeat and make unhealthy food choices [53]. This multidimensionality and individual nature of hunger cues [59], combined with the effect specific diets can have on sensitivity to interoceptive hunger and satiety cues [112], shows the need for individualized eating interventions which include interoceptive aspects.

### 4.4. Alcohol consumption

In line with previous reviews [113, 114], the current review found that the consumption of alcohol impairs interoceptive accuracy. Alcohol has a negative effect on the functioning of the insular cortex [33, 52, 57, 60, 65, 114], a region of the brain that is important in the processing of interoceptive signals. Additionally, the insular cortex plays a key role in decision-making processes. These processes rely on emotional signals from the body, signals that individuals with substance use disorders are less likely to monitor effectively. Damage to the insular cortex, for example caused by alcohol consumption, can result in impaired interoceptive accuracy [115]. Subsequently, non-emotional sources (such as habits) are used to manage decision-making processes, leading to the making of more disadvantageous choices [33, 52, 57, 60]. Interestingly, while interoceptive accuracy is impaired by the use of alcohol, some studies found that interoceptive attention increases [45, 74]. The increase in interoceptive attention may be a compensatory strategy for poor interoceptive accuracy. Previous and current findings indicate the acute and long-term negative influence of alcohol use on interoception, while better interoceptive skills may prevent alcohol abuse.

### 4.5. Exercise

Our results indicate that acute exercise has a positive influence on self-reported interoceptive attention. One possibility for the increase in interoceptive attention during exercise, is that increased arousal narrows attentional resources on physiological signals [64, 104]. Predictive coding models say that predictions are compared against afferent information to produce error signals, which are then used for interpretation of incoming stimuli and ensuring the correct behavioral responses [10]. The utilization of attentional resources can be trained through exercise interventions focusing on interoception, resulting in increases in exercise tolerance, self-control, and confidence and trust in physical abilities [66, 81]. On the other hand, results regarding interoceptive accuracy identified in the current review were conflicting. Studies indicating better interoceptive accuracy during acute exercise show an increase in anterior cruciate cortex and insular cortex activity [65, 67], which is in line with a review article that additionally mentions an increase in neurochemical activity during exercise to explain the role of interoception in exercise [10]. Exercise background or experience influences baseline interoceptive accuracy [104], allowing for more finely tuned self-regulation of physical needs during exercise [95]. Conversely, two studies indicate that interoceptive accuracy does not influence physical performance [96, 98], while another study adds that interoceptive accuracy at rest is only transferable to very low physical loads [97]. This latter study explains that this might be the case because of higher internal noise (e.g., increased overall arousal, faster breathing, or rhythmic movements) during exercise. Also, physical, physiological, and interoceptive responses to exercise might be influenced by the intensity of the exercise, whether the exercise is dynamic or static, and the design of the exercise [98]. Interestingly, only one article investigated the longitudinal effects of exercise on interoception, where most studies focused on the effect of acute exercise on interoceptive abilities. This longitudinal study found that interoception only increased after two to three months of moderate intensity, indicating that repeated exposure to exercise may be necessary for exercise to influence interoceptive abilities [67]. Moreover, the studies in this section focused on cardiovascular exercise, thus we cannot say anything about the relation between interoception and other forms of exercise (e.g., resistance training).

### 4.6. Smoking

Few studies on the relation between interoception and smoking were found, indicating a lack of research on this relation. The only study investigating differences between smokers and non-smokers showed that non-smokers performed better on an interoceptive accuracy task, while there were no differences in interoceptive attention [43]. It was argued that specific brain areas involved with interoceptive accuracy are affected by cigarette smoking, while areas associated with subjective interpretation of bodily signals remain intact. Another study adds to this by showing that insula activity is decreased in cigarette smokers during nicotine withdrawal, along with changes in physiological and behavioral interoceptive measures [34]. Comparable to the line of reasoning in the alcohol consumption section, complex decision-making processes are reliant upon bodily signals, which are processed in the ventromedial prefrontal cortex, insula, and amygdala. Structural and functional damage to these regions may cause substance addicts to be less sensitive to internal signals about negative consequences of drug use, increasing engagement in risky behaviors [115]. However, mindfulness based interventions may be an effective tool to improve interoceptive abilities and lessen the severity of substance use disorders (including alcohol use and cigarette smoking) [77, 78]. Unlike the studies on cigarette smoking in this review, research in individuals using smoked or crack cocaine seems to indicate an increase in interoceptive processing as a result of drug use, but this may be dependent on the type of drug or the way the drug is administered [38]. While the use of cigarettes seems to negatively impact interoception, research on the relation between smoking and interoception should be expanded to draw conclusions.

### 4.7. Sleep

While the current findings on the interaction between interoception and sleep did not allow for strong conclusions, the identified studies may shed light on the way the body reacts to poor sleep quality and stressful situations. One study found that during the COVID-19 lockdown, greater interoceptive attention was linked to greater subjective well-being (including sleep quality) [62]. This study also notes that individual differences in interoception are associated with physiological health, through heart rate variability (HRV). HRV is not only linked to interoception but is also a biomarker of stress and can be an indicator of resilience under stressful situations [62]. This indicates the crucial relation between interoception, stress, and general well-being (including sleep quality). Impairments in interoceptive attention may alter psychological and neurological factors, which leads to changes in mood and sleep quality [32]. Conversely, interoceptive accuracy was found to be increased in individuals with greater sleep difficulties [39]. One explanation for the finding that poor sleep quality relates to decreased interoceptive attention and increased interoceptive accuracy may be because interoception during sleep is related to the heartbeat evoked cortical potential representing cognitive information processing [36]. Research on sleep deprivation has shown sleep-deprived individuals are able to accurately assess cognitive task performance [116], and sleep-deprived doctors are able to monitor work performance successfully [117]. It may be possible that good sleep quality improves interoception, but when sleep quality is impaired interoceptive pathways ensure proper cognitive function.

### 4.8. Strengths and limitations

To the best of our knowledge, this is the first review to provide a comprehensive overview of the relation between interoception and several lifestyle factors. The overarching view of this review is a strength, as it makes it possible to cross-compare the role of interoception between different lifestyle factors. This allows for the identification of lifestyle factors where the role of interoception has been investigated most extensively, as opposed to those lifestyle factors where there is a lack of research. Additionally, by synthesizing findings per lifestyle factor, generalizable conclusions can be made to support evidence-based decision-making processes in a clear and structured manner.

We used the 2×2 factorial structure for interoception to synthesize results found in the included studies. The model constantly asks two questions which are easy to answer (“what was measured?” and “how was this measured?”), and thus ensures quality of subdivision of studies and creates a complete picture of interoception. While we view the use of this model as a strength of this review, it was limited by studies that only included one of the quadrants from the 2×2 model (e.g., accuracy and objective performance), presenting only part of the complex nature of interoception. Also, since this model was developed only recently, many articles included here did not use the exact same terms used in the 2×2 structure. Many studies on interoception use the same terms for different domains, or different terms for the same domains [118]. This means we had to make choices as to the placement of the studies (or parts of studies) within the 2×2 factorial structure, which could subsequently lead to studies being placed within an interoceptive domain the authors of the study did not intend to investigate. However, we are confident in the correct placement of individual studies, as the model is easy to interpret. In fact, we would argue for increased use of this model, as it creates clarity in the definitions of interoceptive domains and allows for cross-correlations between domains and measures (e.g., the correlation between objective performance tests and self-reported beliefs, which is defined as interoceptive awareness).

Based on literature, we determined six overarching lifestyle factors, which were written out in detail in the search strings. While this allowed for an overview and cross-comparisons of the relation between interoception and lifestyle factors, it might have resulted in studies not being found. For example, in the cognitive leisure and relaxation activities part of the search string, terms like leisure activities, cognitive stimulation, and mental stimulation were used. This resulted in identifying several articles on mindfulness, tai-chi, yoga, and meditation, even though these terms were not specifically mentioned in the search strings. This choice was made to cover studies investigating the role of cognitive leisure activities such as reading, gardening, or puzzles in interoception, as well as the mindfulness-related activities. Nevertheless, the current findings give a good overview of the influence of mindfulness on interoceptive abilities, while also showing the lack of research on other cognitive stimulating activities. Future review articles specifically focusing on the role of mindfulness, tai-chi, yoga, and meditation in interoception, could define these constructs more comprehensively in their search strings.

### 4.9. Implications for practical and future research

This review showed that mindfulness, and practices related to mindfulness, seem to be effective in improving interoceptive skills and may therefore be recommended to be included in lifestyle interventions. For example, mindfulness-based interventions targeting individuals suffering from substance use disorders have been shown to be beneficial in improving interoception and decreasing substance related problems [77, 78]. Furthermore, interventions aimed at improving intuitive eating behavior should also consider adding interoceptive training, as this review indicates that intuitive eating and interoception are closely related.

Research investigating the relation between interoception and exercise are currently mainly focusing on the effect of acute exercise on interoception. Additional research should also consider the effect of longitudinal exposure to exercise on interoception, or vice versa. Consistently participating in exercise is beneficial for long-term health, thus understanding the role of interoception can help in the development of longitudinal exercise programs. Additionally, specific types of exercise might influence the development of interoceptive abilities differently, and interoception might be more beneficial in some exercise activities compared to others. Understanding which exercise types promote interoception the most, and vice versa, can help in the development of lifestyle interventions.

The 2×2 factorial structure for interoception reveals two literature gaps. First, there’s a lack of measures for objective interoceptive attention, suggesting the need for new instruments like experience-sampling methods. Second, interoceptive awareness, defined as the relationship between performance and beliefs, is rarely quantified. Only 6 out of 21 studies that used both performance and belief measures did so. Future research should examine at least two quadrants of the 2×2 structure to increase our understanding of the different interoception domains.

## 5. Conclusion

The current review indicates the importance of interoception in various health behaviors. Strong support was found indicating the importance of mindfulness qualities and interventions in the promotion of interoception. The evidence for a positive relation between interoception, eating behavior, and exercise, and the negative relation between interoception and alcohol consumption, was less strong. A limited number of studies of good methodological quality with varying findings were identified on the relation between interoception, smoking behavior, and sleep to draw clear conclusions, which was a gap in literature identified in this review. Moreover, a complete picture of the complex nature of interoception in a healthy lifestyle is not always presented, as indicated by the lack of quantification of interoceptive awareness in the current literature. Thus, future longitudinal studies with more objective measures of interoception are needed in order to better understand the interrelations between interoception and lifestyle factors.

## 6. Other information

### 6.2. Author contributions

Study conception and design was performed by JM, JK. Data collection and analyses were performed by JM, MBo, LV, MBr. The first draft of the manuscript was written by JM. All authors critically revised and commented on previous versions of the manuscript. All authors read and approved the final manuscript.

### 6.4. Availability of data, code, and other materials

The datasets generated and/or analyzed during the current study are available from the corresponding author on reasonable request.

## 9. Appendix

## Appendix A Search String

### PubMed

(Interoception[MeSH] OR interoception[tiab] OR alliesthesia*[tiab] OR “interoceptive accuracy”[tiab] OR “interoceptive sensibilit*”[tiab] OR “interoceptive sensitivit*”[tiab] OR “interoceptive awareness”[tiab] OR “interoceptive attention”[tiab]) AND (((((((Smoking[MeSH] OR smoking[tiab] OR “smoking behavior*”[tiab] OR “smoking habit*”[tiab] OR smokers[MeSH] OR smoker*[tiab] OR “tobacco products”[MeSH] OR “tobacco product*”[tiab] OR cigar*[tiab] OR “tobacco use”[MeSH] OR “tobacco use*”[tiab] OR “tobacco consumption”[tiab] OR “tobacco chewing”[tiab] OR “tobacco dependence”[tiab] OR “tobacco use cessation”[MeSH] OR “tobacco use cessation*”[tiab] OR “tobacco cessation*”[tiab] OR “tobacco use disorder”[MeSH] OR “tobacco use disorder*”[tiab] OR “tobacco-use disorder*”[tiab] OR “nicotine addiction*”[tiab] OR “nicotine use disorder*”[tiab] OR “nicotine dependence”[tiab] OR “tobacco smoking”[MeSH] OR “tobacco smoking”[tiab]) OR (”Life style”[MeSH] OR “life style*”[tiab] OR lifestyle*[tiab] OR “life style induced illness”[tiab] OR “lifestyle factor*”[tiab] OR “healthy lifestyle”[MeSH] or “healthy lifestyle*”[tiab] OR “healthy life style*”[tiab])) OR (”Diet, healthy”[MeSH] OR “healthy diet*”[tiab] OR “healthy eating”[tiab] OR “healthy nutrition”[tiab] OR “prudent diet*”[tiab] OR “feeding behavior”[MeSH] OR “feeding behavior*”[tiab] OR “eating behavior*”[tiab] OR “feeding-related behavior*”[tiab] OR “food habit*”[tiab] OR “eating habit*”[tiab] OR “diet habit*”[tiab] OR “feeding pattern*”[tiab] OR “nutritional status”[MeSH] OR “nutrition* status”[tiab] OR eating[MeSH] OR eating[tiab] OR “food intake”[tiab] OR “feed intake”[tiab] OR “macronutrient intake*”[tiab] OR “micronutrient intake*”[tiab] OR “dietary intake*”[tiab] OR ingestion[tiab] OR “nutrient intake*”[tiab] OR “nutrients intake*”[tiab] OR “feeding and eating disorders”[MeSH] OR “feeding and eating disorder*”[tiab] OR “eating and feeding disorder*”[tiab] OR “feeding disorder*”[tiab] OR “eating disorder*”[tiab] OR “appetite disorder*”[tiab])) OR (”alcohol-induced disorders”[MeSH] OR “alcohol-induced disorder*”[tiab] OR “alcohol-related disorders”[MeSH] OR “alcohol-related disorder*”[tiab] OR “alcohol drinking”[MeSH] OR “alcohol drinking”[tiab] OR alcoholism[MeSH] OR alcoholism[tiab] OR “alcohol dependence”[tiab] OR “alcohol addiction”[tiab] OR “alcohol abuse”[tiab] OR “alcohol use disorder*”[tiab])) OR (”sedentary behavior”[MeSH] OR “sedentary behavior*”[tiab] OR “sedentary lifestyle”[tiab] OR “physical inactivity”[tiab] OR “lack of physical activity”[tiab] OR

exercise[MeSH] OR exercise*[tiab] OR “physical activit*”[tiab] OR “physical exercise*”[tiab] OR “exercise training*”[tiab] OR “aerobic exercise*”[tiab] OR “isometric exercise*”[tiab] OR “physical fitness”[MeSH] OR “physical fitness”[tiab] OR “cardiorespiratory fitness”[MeSH] OR “cardiorespiratory fitness”[tiab] OR “exercise behavior*”[tiab] OR “exercise habit*”[tiab])) OR (”leisure activities”[MeSH] OR “leisure activit*”[tiab] OR leisure*[tiab] OR “cognitive leisure activit*”[tiab] OR “cognitive stimulation”[tiab] OR “mental stimulation”[tiab] OR cognition[MeSH] OR cognition[tiab] OR “cognitive function*” [tiab])) OR (sleep[MeSH] OR sleep*[tiab] OR “sleep habit*” [tiab] OR “sleeping habit*” [tiab]))

1239 results, 21-09-23

### Embase

exp interoception/ or interoception.ti,ab,cl,oa,kw,kf. or alliesthesia*.ti,ab,cl,oa,kw,kf. or “interoceptive accuracy”.ti,ab,cl,oa,kw,kf. or “interoceptive sensibilit*”.ti,ab,cl,oa,kw,kf. or “interoceptive sensitivit*”.ti,ab,cl,oa,kw,kf. or “interoceptive awareness”.ti,ab,cl,oa,kw,kf. or “interoceptive attention”.ti,ab,cl,oa,kw,kf. AND (exp “life style”/ or “life style*”.ti,ab,cl,oa,kw,kf. or lifestyle*.ti,ab,cl,oa,kw,kf. or “life style induced illness”.ti,ab,cl,oa,kw,kf. or “lifestyle factor*”.ti,ab,cl,oa,kw,kf. or exp “healthy lifestyle”/ or “healthy lifestyle*”.ti,ab,cl,oa,kw,kf. or “healthy life style*”.ti,ab,cl,oa,kw,kf. OR exp “diet, healthy”/ or “healthy diet*”.ti,ab,cl,oa,kw,kf. or “healthy eating”.ti,ab,cl,oa,kw,kf. or “healthy nutrition”.ti,ab,cl,oa,kw,kf. or “prudent diet*”.ti,ab,cl,oa,kw,kf. or exp “feeding behavior”/ or “feeding behavior*”.ti,ab,cl,oa,kw,kf. or “eating behavior*”.ti,ab,cl,oa,kw,kf. or “feeding-related behavior*”.ti,ab,cl,oa,kw,kf. or “food habit*”.ti,ab,cl,oa,kw,kf. or “eating habit*”.ti,ab,cl,oa,kw,kf. or “diet habit*”.ti,ab,cl,oa,kw,kf. or “feeding pattern*”.ti,ab,cl,oa,kw,kf. or exp “nutritional status”/ or “nutrition* status”.ti,ab,cl,oa,kw,kf. or exp eating/ or eating.ti,ab,cl,oa,kw,kf. or “food intake”.ti,ab,cl,oa,kw,kf. or “feed intake”.ti,ab,cl,oa,kw,kf. or “macronutrient intake*”.ti,ab,cl,oa,kw,kf. or “micronutrient intake*”.ti,ab,cl,oa,kw,kf. or “dietary intake*”.ti,ab,cl,oa,kw,kf. or ingestion.ti,ab,cl,oa,kw,kf. or “nutrient intake*”.ti,ab,cl,oa,kw,kf. or “nutrients intake*”.ti,ab,cl,oa,kw,kf. or “feeding and exp eating disorders”/ or “feeding and eating disorder*”.ti,ab,cl,oa,kw,kf. or “eating and feeding disorder*”.ti,ab,cl,oa,kw,kf. or “feeding disorder*”.ti,ab,cl,oa,kw,kf. or “eating disorder*”.ti,ab,cl,oa,kw,kf. or “appetite disorder*”.ti,ab,cl,oa,kw,kf. OR exp “alcohol-induced disorders”/ or “alcohol-induced disorder*”.ti,ab,cl,oa,kw,kf. or exp “alcohol-related disorders”/ or “alcohol-related disorder*”.ti,ab,cl,oa,kw,kf. or exp “alcohol drinking”/ or “alcohol drinking”.ti,ab,cl,oa,kw,kf. or exp alcoholism/ or alcoholism.ti,ab,cl,oa,kw,kf. or “alcohol dependence”.ti,ab,cl,oa,kw,kf. or “alcohol addiction”.ti,ab,cl,oa,kw,kf. or “alcohol abuse”.ti,ab,cl,oa,kw,kf. or “alcohol use disorder*”.ti,ab,cl,oa,kw,kf. OR exp “sedentary behavior”/ or “sedentary behavior*”.ti,ab,cl,oa,kw,kf. or “sedentary lifestyle”.ti,ab,cl,oa,kw,kf. or “physical inactivity”.ti,ab,cl,oa,kw,kf. or “lack of physical activity”.ti,ab,cl,oa,kw,kf. or exp exercise/ or exercise*.ti,ab,cl,oa,kw,kf. or “physical activit*”.ti,ab,cl,oa,kw,kf. or “physical exercise*”.ti,ab,cl,oa,kw,kf. or “exercise training*”.ti,ab,cl,oa,kw,kf. or “aerobic exercise*”.ti,ab,cl,oa,kw,kf. or “isometric exercise*”.ti,ab,cl,oa,kw,kf. or exp “physical fitness”/ or “physical fitness”.ti,ab,cl,oa,kw,kf. or exp “cardiorespiratory fitness”/ or “cardiorespiratory fitness”.ti,ab,cl,oa,kw,kf. or “exercise behavior*”.ti,ab,cl,oa,kw,kf. or “exercise habit*”.ti,ab,cl,oa,kw,kf. OR exp “leisure activities”/ or “leisure activit*”.ti,ab,cl,oa,kw,kf. or leisure*.ti,ab,cl,oa,kw,kf. or “cognitive leisure activit*”.ti,ab,cl,oa,kw,kf. or “cognitive stimulation”.ti,ab,cl,oa,kw,kf. or “mental stimulation”.ti,ab,cl,oa,kw,kf. or exp cognition/ or cognition.ti,ab,cl,oa,kw,kf. or “cognitive function*”.ti,ab,cl,oa,kw,kf. OR exp sleep/ or sleep*.ti,ab,cl,oa,kw,kf. or “sleep habit*”.ti,ab,cl,oa,kw,kf. or “sleeping habit*”.ti,ab,cl,oa,kw,kf. OR exp smoking/ or smoking.ti,ab,cl,oa,kw,kf. or “smoking behavior*”.ti,ab,cl,oa,kw,kf. or “smoking habit*”.ti,ab,cl,oa,kw,kf. or exp smokers/ or smoker*.ti,ab,cl,oa,kw,kf. or exp “tobacco products”/ or “tobacco product*”.ti,ab,cl,oa,kw,kf. or cigar*.ti,ab,cl,oa,kw,kf. or exp “tobacco use”/ or “tobacco use*”.ti,ab,cl,oa,kw,kf. or “tobacco consumption”.ti,ab,cl,oa,kw,kf. or “tobacco chewing”.ti,ab,cl,oa,kw,kf. or “tobacco dependence”.ti,ab,cl,oa,kw,kf. or exp “tobacco use cessation”/ or “tobacco use cessation*”.ti,ab,cl,oa,kw,kf. or “tobacco cessation*”.ti,ab,cl,oa,kw,kf. or exp “tobacco use disorder”/ or “tobacco use disorder*”.ti,ab,cl,oa,kw,kf. or “tobacco-use disorder*”.ti,ab,cl,oa,kw,kf. or “nicotine addiction*”.ti,ab,cl,oa,kw,kf. or “nicotine use disorder*”.ti,ab,cl,oa,kw,kf. or “nicotine dependence”.ti,ab,cl,oa,kw,kf. or exp “tobacco smoking”/ or “tobacco smoking”.ti,ab,cl,oa,kw,kf.)

2759 results, 21-09-23

### Web of Science

(TS=(Interoception alliesthesia* OR “interoceptive accuracy” OR “interoceptive sensibilit*” OR “interoceptive sensitivit*” OR “interoceptive awareness” OR “interoceptive attention”)) AND (TS=(Smoking OR “smoking behavior*” OR “smoking habit*” OR smokers OR smoker* OR “tobacco products” OR “tobacco product*” OR cigar* OR “tobacco use” OR “tobacco use*” OR “tobacco consumption” OR “tobacco chewing” OR “tobacco dependence” OR “tobacco use cessation” OR “tobacco use cessation*” OR “tobacco cessation*” OR “tobacco use disorder” OR “tobacco use disorder*” OR “tobacco-use disorder*” OR “nicotine addiction*” OR “nicotine use disorder*” OR “nicotine dependence” OR “tobacco smoking” OR “tobacco smoking”) OR TS=(”Life style” OR “life style*” OR lifestyle* OR “life style induced illness” OR “lifestyle factor*” OR “healthy lifestyle” or “healthy lifestyle*” OR “healthy life style*”) OR TS=(”Diet, healthy” OR “healthy diet*” OR “healthy eating” OR “healthy nutrition” OR “prudent diet*” OR “feeding behavior” OR “feeding behavior*” OR “eating behavior*” OR “feeding-related behavior*” OR “food habit*” OR “eating habit*” OR “diet habit*” OR “feeding pattern*” OR “nutritional status” OR “nutrition* status” OR eating OR eating OR “food intake” OR “feed intake” OR “macronutrient intake*” OR “micronutrient intake*” OR “dietary intake*” OR ingestion OR “nutrient intake*” OR “nutrients intake*” OR “feeding and eating disorders” OR “feeding and eating disorder*” OR “eating and feeding disorder*” OR “feeding disorder*” OR “eating disorder*” OR “appetite disorder*”) OR TS=(”alcohol-induced disorders” OR “alcohol-induced disorder*” OR “alcohol-related disorders” OR “alcohol-related disorder*” OR “alcohol drinking” OR “alcohol drinking” OR alcoholism OR alcoholism OR “alcohol dependence” OR “alcohol addiction” OR “alcohol abuse” OR “alcohol use disorder*”) OR TS=(”sedentary behavior” OR “sedentary behavior*” OR “sedentary lifestyle” OR “physical inactivity” OR “lack of physical activity” OR exercise OR exercise* OR “physical activit*” OR “physical exercise*” OR “exercise training*” OR “aerobic exercise*” OR “isometric exercise*” OR “physical fitness” OR “physical fitness” OR “cardiorespiratory fitness” OR “cardiorespiratory fitness” OR “exercise behavior*” OR “exercise habit*”) OR TS=(”leisure activities” OR “leisure activit*” OR leisure* OR “cognitive leisure activit*” OR “cognitive stimulation” OR “mental stimulation” OR cognition OR cognition OR “cognitive function*”) OR TS=(sleep OR sleep* OR “sleep habit*” OR “sleeping habit*”))

611 results, 21-09-23

## Appendix B Quality score

This quality score was used to assess the quality of included studies in this systematic review and is applicable to both interventional and observational studies. The score was designed based on previously published scoring systems. The quality score consists of 5 items, and each item is allocated 0, 1 or 2 points. This allows a total score between 0 and 10 points, 10 representing the highest quality.

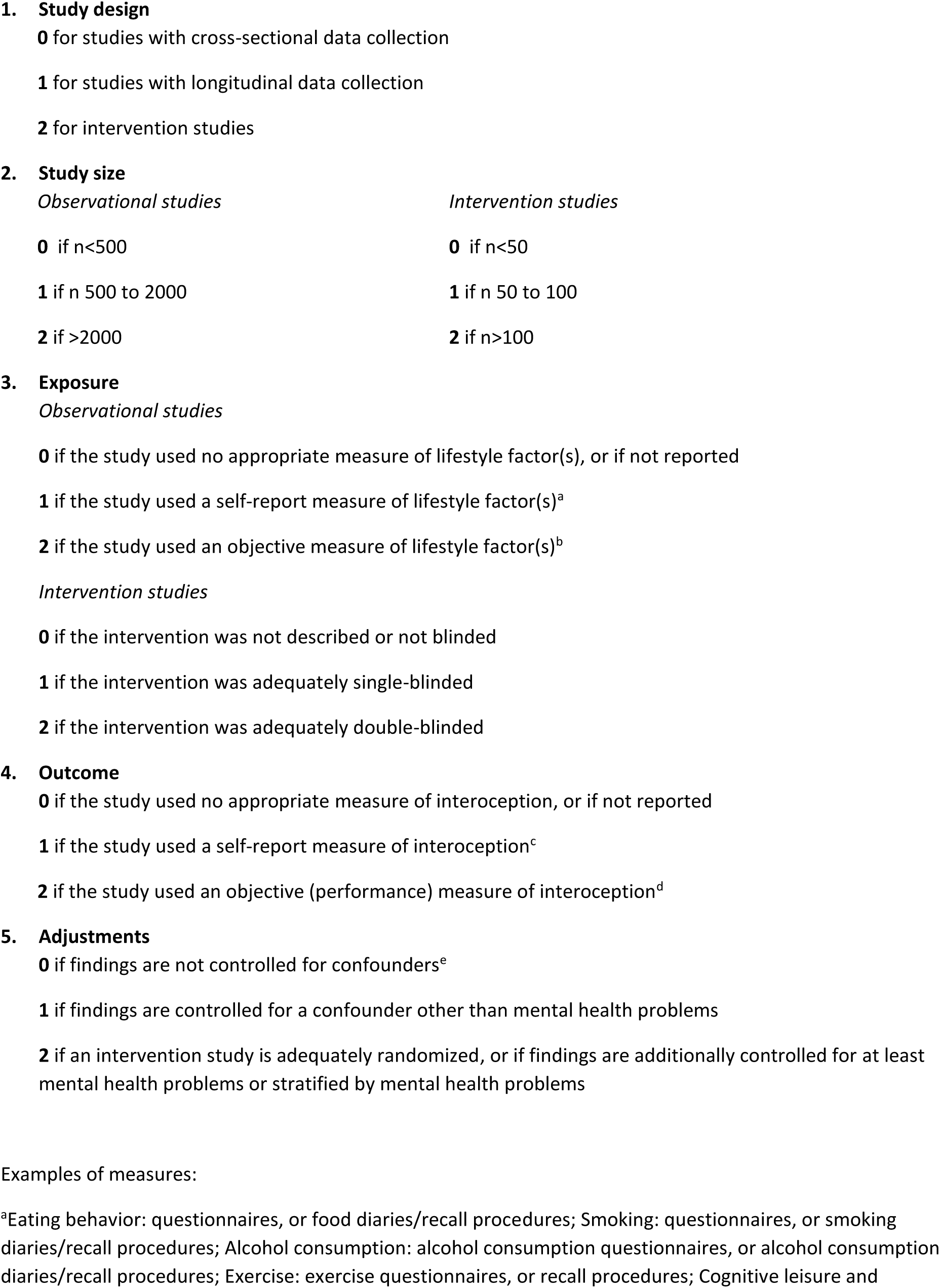
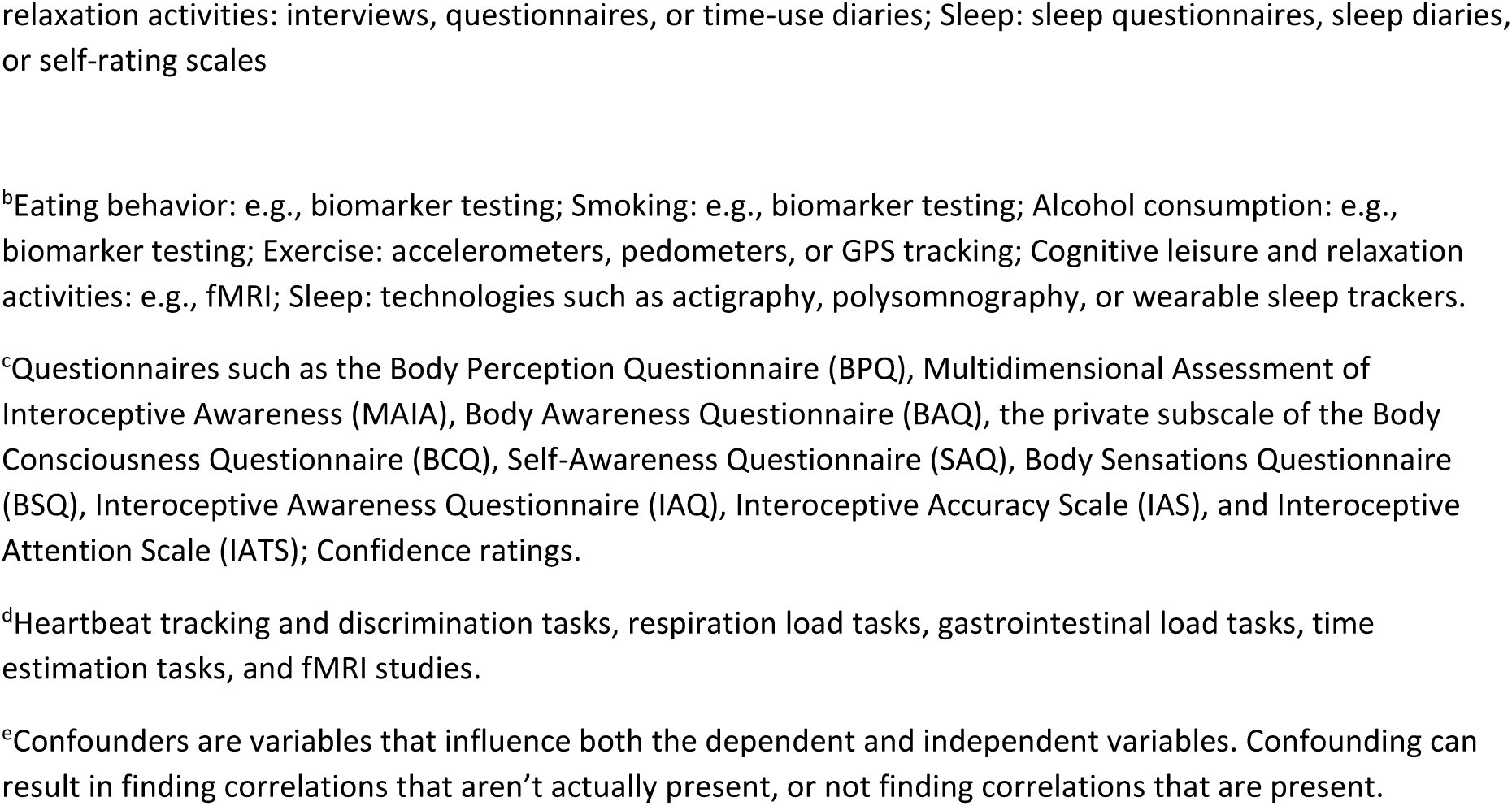

## References

1. World Health Organization. Noncommunicable diseases 2023 [updated September 16, 2023. Available from: https://www.who.int/news-room/fact-sheets/detail/noncommunicable-diseases.

2. Harris KM, McDade TW. The Biosocial Approach to Human Development, Behavior, and Health Across the Life Course. RSF. 2018;4(4):2–26. 10.7758/RSF.2018.4.4.01.

3. Murphy J, Catmur C, Bird G. Classifying individual differences in interoception: Implications for the measurement of interoceptive awareness. Psychon Bull Rev. 2019;26(5):1467–71. 10.3758/s13423-019-01632-7.

4. Farb N, Daubenmier J, Price CJ, Gard T, Kerr C, Dunn BD, et al. Interoception, contemplative practice, and health. Front Psychol. 2015;6:763. 10.3389/fpsyg.2015.00763.

5. Quadt L, Critchley HD, Garfinkel SN. The neurobiology of interoception in health and disease. Ann N Y Acad Sci. 2018;1428(1):112–28. 10.1111/nyas.13915.

6. Critchley HD, Harrison NA. Visceral influences on brain and behavior. Neuron. 2013;77(4):624–38. 10.1016/j.neuron.2013.02.008.

7. Dantzer R, Kelley KW. Twenty years of research on cytokine-induced sickness behavior. Brain Behav Immun. 2007;21(2):153–60. 10.1016/j.bbi.2006.09.006.

8. Tracey KJ. Reflex control of immunity. Nat Rev Immunol. 2009;9(6):418–28. 10.1038/nri2566.

9. Dantzer R, O’Connor JC, Freund GG, Johnson RW, Kelley KW. From inflammation to sickness and depression: when the immune system subjugates the brain. Nat Rev Neurosci. 2008;9(1):46–56. 10.1038/nrn2297.

10. Wallman-Jones A, Perakakis P, Tsakiris M, Schmidt M. Physical activity and interoceptive processing: Theoretical considerations for future research. Int J Psychophysiol. 2021;166:38–49. 10.1016/j.ijpsycho.2021.05.002.

11. Bajpai G, Nahrendorf M. Infectious and lifestyle modifiers of immunity and host resilience. Immunity. 2021;54(6):1110–22. 10.1016/j.immuni.2021.05.011.

12. Monye I, Adelowo AB. Strengthening immunity through healthy lifestyle practices: Recommendations for lifestyle interventions in the management of COVID-19. Lifestyle Med (Hoboken). 2020;1(1):e7. 10.1002/lim2.7.

13. Garfinkel SN, Tiley C, O’Keeffe S, Harrison NA, Seth AK, Critchley HD. Discrepancies between dimensions of interoception in autism: Implications for emotion and anxiety. Biol Psychol. 2016;114:117–26. 10.1016/j.biopsycho.2015.12.003.

14. Shah P, Hall R, Catmur C, Bird G. Alexithymia, not autism, is associated with impaired interoception. Cortex. 2016;81:215–20. 10.1016/j.cortex.2016.03.021.

15. Page MJ, McKenzie JE, Bossuyt PM, Boutron I, Hoffmann TC, Mulrow CD, et al. The PRISMA 2020 statement: an updated guideline for reporting systematic reviews. BMJ. 2021;372:n71. 10.1136/bmj.n71.

16. Martin E, Dourish CT, Rotshtein P, Spetter MS, Higgs S. Interoception and disordered eating: A systematic review. Neurosci Biobehav Rev. 2019;107:166–91. 10.1016/j.neubiorev.2019.08.020.

17. The EndNote Team. EndNote. EndNote 21 ed. Philadelphia, PA: Clarivate; 2013.

18. van de Schoot R, de Bruin J, Schram R, Zahedi P, de Boer J, Weijdema F, et al. An open source machine learning framework for efficient and transparent systematic reviews. Nat Mach Intell. 2021;3(2):125–33. 10.1038/s42256-020-00287-7.

19. Ferdinands G. AI-Assisted Systematic Reviewing: Selecting Studies to Compare Bayesian Versus Frequentist SEM for Small Sample Sizes. Multivariate Behav Res. 2021;56(1):153–4. 10.1080/00273171.2020.1853501.

20. Collins SE, Kirouac M. Alcohol Consumption. In: Gellman MD, Turner JR, editors. Encyclopedia of Behavioral Medicine. New York, NY: Springer New York; 2013. p. 61–5.

21. Malinowski P, Shalamanova L. Meditation and Cognitive Ageing: the Role of Mindfulness Meditation in Building Cognitive Reserve. J Cogn Enhance. 2017;1(2):96–106. 10.1007/s41465-017-0022-7.

22. Saczynski JS, Jonsdottir MK, Sigurdsson S, Eiriksdottir G, Jonsson PV, Garcia ME, et al. White matter lesions and cognitive performance: the role of cognitively complex leisure activity. J Gerontol A Biol Sci Med Sci. 2008;63(8):848–54. 10.1093/gerona/63.8.848.

23. Verghese J, LeValley A, Derby C, Kuslansky G, Katz M, Hall C, et al. Leisure activities and the risk of amnestic mild cognitive impairment in the elderly. Neurology. 2006;66(6):821–7. 10.1212/01.wnl.0000202520.68987.48.

24. Wilson RS, Barnes LL, Aggarwal NT, Boyle PA, Hebert LE, Mendes de Leon CF, et al. Cognitive activity and the cognitive morbidity of Alzheimer disease. Neurology. 2010;75(11):990–6. 10.1212/WNL.0b013e3181f25b5e.

25. LaCaille L, Patino-Fernandez AM, Monaco J, Ding D, Upchurch Sweeney CR, Butler CD, et al. Eating Behavior. In: Gellman MD, Turner JR, editors. Encyclopedia of Behavioral Medicine. New York, NY: Springer New York; 2013. p. 641–2.

26. Caspersen CJ, Powell KE, Christenson GM. Physical activity, exercise, and physical fitness: definitions and distinctions for health-related research. Public Health Rep. 1985;100(2):126–31.

27. Baranwal N, Yu PK, Siegel NS. Sleep physiology, pathophysiology, and sleep hygiene. Prog Cardiovasc Dis. 2023;77:59–69. 10.1016/j.pcad.2023.02.005.

28. Baker E, Webb Hooper M. Smoking Behavior. In: Gellman MD, Turner JR, editors. Encyclopedia of Behavioral Medicine. New York, NY: Springer New York; 2013. p. 1818–20.

29. Stroup DF, Berlin JA, Morton SC, Olkin I, Williamson GD, Rennie D, et al. Meta-analysis of observational studies in epidemiology: a proposal for reporting. Meta-analysis Of Observational Studies in Epidemiology (MOOSE) group. JAMA. 2000;283(15):2008–12. 10.1001/jama.283.15.2008.

30. von Elm E, Altman DG, Egger M, Pocock SJ, Gotzsche PC, Vandenbroucke JP, et al. The Strengthening the Reporting of Observational Studies in Epidemiology (STROBE) Statement: guidelines for reporting observational studies. Int J Surg. 2014;12(12):1495–9. 10.1016/j.ijsu.2014.07.013.

31. Wong WC, Cheung CS, Hart GJ. Development of a quality assessment tool for systematic reviews of observational studies (QATSO) of HIV prevalence in men having sex with men and associated risk behaviours. Emerg Themes Epidemiol. 2008;5:23. 10.1186/1742-7622-5-23.

32. Arora T, Barbato M, Al Hemeiri S, Omar OM, AlJassmi MA. A mysterious sensation about sleep and health: the role of interoception. BMC Public Health. 2021;21(1):1584. 10.1186/s12889-021-11603-0.

33. Ates Col I, Sonmez MB, Vardar ME. Evaluation of Interoceptive Awareness in Alcohol-Addicted Patients. Noro Psikiyatr Ars. 2016;53(1):17–22. 10.5152/npa.2015.9898.

34. Avery JA, Burrows K, Kerr KL, Bodurka J, Khalsa SS, Paulus MP, et al. How the Brain Wants What the Body Needs: The Neural Basis of Positive Alliesthesia. Neuropsychopharmacology. 2017;42(4):822–30. 10.1038/npp.2016.128.

35. Avery JA, Powell JN, Breslin FJ, Lepping RJ, Martin LE, Patrician TM, et al. Obesity is associated with altered mid-insula functional connectivity to limbic regions underlying appetitive responses to foods. J Psychopharmacol. 2017;31(11):1475–84. 10.1177/0269881117728429.

36. Billeci L, Faraguna U, Santarcangelo EL, d’Ascanio P, Varanini M, Sebastiani L. Heartbeat-Evoked Cortical Potential during Sleep and Interoceptive Sensitivity: A Matter of Hypnotizability. Brain Sci. 2021;11(8). 10.3390/brainsci11081089.

37. Daubenmier J, Sze J, Kerr CE, Kemeny ME, Mehling W. Follow your breath: respiratory interoceptive accuracy in experienced meditators. Psychophysiology. 2013;50(8):777–89. 10.1111/psyp.12057.

38. de la Fuente A, Sedeno L, Vignaga SS, Ellmann C, Sonzogni S, Belluscio L, et al. Multimodal neurocognitive markers of interoceptive tuning in smoked cocaine. Neuropsychopharmacology. 2019;44(8):1425–34. 10.1038/s41386-019-0370-3.

39. Ewing DL, Manassei M, Gould van Praag C, Philippides AO, Critchley HD, Garfinkel SN. Sleep and the heart: Interoceptive differences linked to poor experiential sleep quality in anxiety and depression. Biol Psychol. 2017;127:163–72. 10.1016/j.biopsycho.2017.05.011.

40. Hanley AW, Mehling WE, Garland EL. Holding the body in mind: Interoceptive awareness, dispositional mindfulness and psychological well-being. J Psychosom Res. 2017;99:13–20. 10.1016/j.jpsychores.2017.05.014.

41. Herbert BM, Blechert J, Hautzinger M, Matthias E, Herbert C. Intuitive eating is associated with interoceptive sensitivity. Effects on body mass index. Appetite. 2013;70:22–30. 10.1016/j.appet.2013.06.082.

42. Herbert BM, Pollatos O. Attenuated interoceptive sensitivity in overweight and obese individuals. Eat Behav. 2014;15(3):445–8. 10.1016/j.eatbeh.2014.06.002.

43. Hina F, Aspell JE. Altered interoceptive processing in smokers: Evidence from the heartbeat tracking task. Int J Psychophysiol. 2019;142:10–6. 10.1016/j.ijpsycho.2019.05.012.

44. Iatridi V, Quadt L, Hayes JE, Garfinkel SN, Yeomans MR. Female sweet-likers have enhanced cross-modal interoceptive abilities. Appetite. 2021;165:105290. 10.1016/j.appet.2021.105290.

45. Jakubczyk A, Skrzeszewski J, Trucco EM, Suszek H, Zaorska J, Nowakowska M, et al. Interoceptive accuracy and interoceptive sensibility in individuals with alcohol use disorder-Different phenomena with different clinical correlations? Drug Alcohol Depend. 2019;198:34–8. 10.1016/j.drugalcdep.2019.01.036.

46. Khalsa SS, Rudrauf D, Damasio AR, Davidson RJ, Lutz A, Tranel D. Interoceptive awareness in experienced meditators. Psychophysiology. 2008;45(4):671–7. 10.1111/j.1469-8986.2008.00666.x.

47. Khalsa SS, Rudrauf D, Hassanpour MS, Davidson RJ, Tranel D. The practice of meditation is not associated with improved interoceptive awareness of the heartbeat. Psychophysiology. 2019;57(2):e13479. 10.1111/psyp.13479.

48. Kiken LG, Shook NJ, Robins JL, Clore JN. Association between mindfulness and interoceptive accuracy in patients with diabetes: Preliminary evidence from blood glucose estimates. Complement Ther Med. 2018;36:90–2. 10.1016/j.ctim.2017.12.003.

49. Lovan P, Prado G, Lee T, Coccia C. A snapshot of eating behaviors in undergraduate college students living in South Florida. J Am Coll Health. 2022:1–10. 10.1080/07448481.2022.2119402.

50. Mailloux G, Bergeron S, Meilleur D, D’Antono B, Dube I. Examining the associations between overeating, disinhibition, and hunger in a nonclinical sample of college women. Int J Behav Med. 2014;21(2):375–84. 10.1007/s12529-013-9306-1.

51. Melloni M, Sedeno L, Couto B, Reynoso M, Gelormini C, Favaloro R, et al. Preliminary evidence about the effects of meditation on interoceptive sensitivity and social cognition. Behav Brain Funct. 2013;9:47. 10.1186/1744-9081-9-47.

52. Meric I, Sonmez MB. Decision-making, interoceptive awareness and mindful attention awareness in male patients with alcohol use disorder. Cogn Neuropsychiatry. 2022;27(1):35–48. 10.1080/13546805.2021.2011183.

53. Nishimura A, Harashima SI, Hosoda K, Honda I. Overeating Risk in Overweight Young Women Is Divided into Two Types According to Appetite and Eating Behavior. Metab Syndr Relat Disord. 2020;18(9):435–42. 10.1089/met.2020.0042.

54. Oswald A, Chapman J, Wilson C. Do interoceptive awareness and interoceptive responsiveness mediate the relationship between body appreciation and intuitive eating in young women? Appetite. 2017;109:66–72. 10.1016/j.appet.2016.11.019.

55. Price CJ, Crowell SE, Pike KC, Cheng SC, Puzia M, Thompson EA. Psychological and Autonomic Correlates of Emotion Dysregulation among Women in Substance Use Disorder Treatment. Subst Use Misuse. 2019;54(1):110–9. 10.1080/10826084.2018.1508297.

56. Robinson E, Marty L, Higgs S, Jones A. Interoception, eating behaviour and body weight. Physiol Behav. 2021;237:113434. 10.1016/j.physbeh.2021.113434.

57. Schmidt AF, Eulenbruch T, Langer C, Banger M. Interoceptive awareness, tension reduction expectancies and self-reported drinking behavior. Alcohol Alcohol. 2013;48(4):472–7. 10.1093/alcalc/agt024.

58. Sönmez MB, Kiliç EK, Çöl IA, Görgülü Y, Çinar RK. Decreased interoceptive awareness in patients with substance use disorders. Journal of Substance Use. 2017;22(1):60–5. 10.3109/14659891.2016.1143048.

59. Stevenson RJ, Hill BJ, Hughes A, Wright M, Bartlett J, Saluja S, et al. Interoceptive hunger, eating attitudes and beliefs. Front Psychol. 2023;14:1148413. 10.3389/fpsyg.2023.1148413.

60. Subay B, Sonmez MB. Interoceptive Awareness, Decision-Making and Impulsiveness in Male Patients with Alcohol or Opioid Use Disorder. Subst Use Misuse. 2021;56(9):1275–83. 10.1080/10826084.2021.1914108.

61. Tylka TL. Development and psychometric evaluation of a measure of intuitive eating. Journal of Counseling Psychology. 2006;53(2):226–40. 10.1037/0022-0167.53.2.226.

62. Vabba A, Porciello G, Monti A, Panasiti MS, Aglioti SM. A longitudinal study of interoception changes in the times of COVID-19: Effects on psychophysiological health and well-being. Heliyon. 2023;9(4):e14951. 10.1016/j.heliyon.2023.e14951.

63. van Strien T. Ice-cream consumption, tendency toward overeating, and personality. Int J Eat Disord. 2000;28(4):460–4. 10.1002/1098-108x(200012)28:4<460::aid-eat16>3.0.co;2-a.

64. Wallman-Jones A, Nigg C, Benzing V, Schmidt M. Leave the screen: The influence of everyday behaviors on self-reported interoception. Biol Psychol. 2023;181:108600. 10.1016/j.biopsycho.2023.108600.

65. Abrams K, Cieslowski K, Johnson S, Krimmel S, La Rosa GB, Barton K, et al. The effects of alcohol on heartbeat perception: Implications for anxiety. Addict Behav. 2018;79:151–8. 10.1016/j.addbeh.2017.12.023.

66. Almarcha M, Gonzalez I, Balague N, Javierre C. Prescribing or co-designing exercise in healthy adults? Effects on mental health and interoceptive awareness. Front Behav Neurosci. 2022;16:944193. 10.3389/fnbeh.2022.944193.

67. Amaya Y, Abe T, Kanbara K, Shizuma H, Akiyama Y, Fukunaga M. The effect of aerobic exercise on interoception and cognitive function in healthy university students: a non-randomized controlled trial. BMC Sports Sci Med Rehabil. 2021;13(1):99. 10.1186/s13102-021-00332-x.

68. de Lima-Araujo GL, de Sousa Junior GM, Mendes T, Demarzo M, Farb N, Barros de Araujo D, et al. The impact of a brief mindfulness training on interoception: A randomized controlled trial. PLoS One. 2022;17(9):e0273864. 10.1371/journal.pone.0273864.

69. Eusebio J, Forbes B, Sahyoun C, Vago DR, Lazar SW, Farb N. Contemplating movement: A randomized control trial of yoga training for mental health. Mental Health and Physical Activity. 2022;23. 10.1016/j.mhpa.2022.100483.

70. Farb NA, Segal ZV, Anderson AK. Mindfulness meditation training alters cortical representations of interoceptive attention. Soc Cogn Affect Neurosci. 2013;8(1):15–26. 10.1093/scan/nss066.

71. Fazia T, Bubbico F, Berzuini G, Tezza LD, Cortellini C, Bruno S, et al. Mindfulness meditation training in an occupational setting: Effects of a 12-weeks mindfulness-based intervention on wellbeing. Work. 2021;70(4):1089–99. 10.3233/WOR-210510.

72. Haase L, Thom NJ, Shukla A, Davenport PW, Simmons AN, Stanley EA, et al. Mindfulness-based training attenuates insula response to an aversive interoceptive challenge. Soc Cogn Affect Neurosci. 2016;11(1):182–90. 10.1093/scan/nsu042.

73. Leganes-Fonteneau M, Bates ME, Islam S, Buckman JF. Changes in interoception after alcohol administration correlate with expectancies and subjective effects. Addict Biol. 2022;27(1):e13098. 10.1111/adb.13098.

74. Leganes-Fonteneau M, Cheang Y, Lam Y, Garfinkel S, Duka T. Interoceptive awareness is associated with acute alcohol-induced changes in subjective effects. Pharmacol Biochem Behav. 2019;181:69–76. 10.1016/j.pbb.2019.03.007.

75. Loucks EB, Nardi WR, Gutman R, Saadeh FB, Li Y, Vago DR, et al. Mindfulness-Based College: A Stage 1 Randomized Controlled Trial for University Student Well-Being. Psychosom Med. 2021;83(6):602–14. 10.1097/PSY.0000000000000860.

76. Palazzo CC, Leghi BE, Diez-Garcia RW. Food Consciousness Intervention Improves Interoceptive Sensitivity and Expression of Exteroception in Women. Nutrients. 2022;14(3). 10.3390/nu14030450.

77. Price CJ, Thompson EA, Crowell S, Pike K. Longitudinal effects of interoceptive awareness training through mindful awareness in body-oriented therapy (MABT) as an adjunct to women’s substance use disorder treatment: A randomized controlled trial. Drug Alcohol Depend. 2019;198:140–9. 10.1016/j.drugalcdep.2019.02.012.

78. Price CJ, Thompson EA, Crowell SE, Pike K, Cheng SC, Parent S, et al. Immediate effects of interoceptive awareness training through Mindful Awareness in Body-oriented Therapy (MABT) for women in substance use disorder treatment. Subst Abus. 2019;40(1):102–15. 10.1080/08897077.2018.1488335.

79. Roberts RL, Ledermann K, Garland EL. Mindfulness-oriented recovery enhancement improves negative emotion regulation among opioid-treated chronic pain patients by increasing interoceptive awareness. J Psychosom Res. 2021;152:110677.10.1016/j.jpsychores.2021.110677.

80. Schillings C, Schultchen D, Pollatos O. Effects of a Single Yoga Session on Cardiac Interoceptive Accuracy and Emotional Experience. Brain Sci. 2021;11(12). 10.3390/brainsci11121572.

81. Teng HC, Yeh ML, Wang MH. Walking with controlled breathing improves exercise tolerance, anxiety, and quality of life in heart failure patients: A randomized controlled trial. Eur J Cardiovasc Nurs. 2018;17(8):717–27. 10.1177/1474515118778453.

82. Thomas EA, Mijangos JL, Hansen PA, White S, Walker D, Reimers C, et al. Mindfulness-Oriented Recovery Enhancement Restructures Reward Processing and Promotes Interoceptive Awareness in Overweight Cancer Survivors: Mechanistic Results From a Stage 1 Randomized Controlled Trial. Integr Cancer Ther. 2019;18:1534735419855138. 10.1177/1534735419855138.

83. Wu Q, Mao X, Luo W, Fan J, Liu X, Wu Y. Enhanced interoceptive attention mediates the relationship between mindfulness training and the reduction of negative mood. Psychophysiology. 2022;59(4):e13991. 10.1111/psyp.13991.

84. Young HA, Gaylor CM, de Kerckhove D, Watkins H, Benton D. Interoceptive accuracy moderates the response to a glucose load: a test of the predictive coding framework. Proc Biol Sci. 2019;286(1898):20190244. 10.1098/rspb.2019.0244.

85. Betka S, Gould Van Praag C, Paloyelis Y, Bond R, Pfeifer G, Sequeira H, et al. Impact of intranasal oxytocin on interoceptive accuracy in alcohol users: an attentional mechanism? Soc Cogn Affect Neurosci. 2018;13(4):440–8. 10.1093/scan/nsy027.

86. Bornemann B, Herbert BM, Mehling WE, Singer T. Differential changes in self-reported aspects of interoceptive awareness through 3 months of contemplative training. Front Psychol. 2014;5:1504. 10.3389/fpsyg.2014.01504.

87. Chen LZ, Dai AY, Yao Y, Si R, Hu Z, Ge L, et al. Effects of 8-Week Tai Chi Chuan Practice on Mindfulness Level. Mindfulness. 2021;12(6):1534–41. 10.1007/s12671-021-01622-8.

88. D’Antoni F, Feruglio S, Matiz A, Cantone D, Crescentini C. Mindfulness Meditation Leads To Increased Dispositional Mindfulness And Interoceptive Awareness Linked To A Reduced Dissociative Tendency. J Trauma Dissociation. 2022;23(1):8–23. 10.1080/15299732.2021.1934935.

89. Fairclough SH, Goodwin L. The effect of psychological stress and relaxation on interoceptive accuracy: Implications for symptom perception. J Psychosom Res. 2007;62(3):289–95. 10.1016/j.jpsychores.2006.10.017.

90. Fischer D, Messner M, Pollatos O. Improvement of Interoceptive Processes after an 8-Week Body Scan Intervention. Front Hum Neurosci. 2017;11:452. 10.3389/fnhum.2017.00452.

91. Flasbeck V, Bamberg C, Brune M. Short-Term Fasting and Ingestion of Caloric Drinks Affect Heartbeat-Evoked Potentials and Autonomic Nervous System Activity in Males. Front Neurosci. 2021;15:622428. 10.3389/fnins.2021.622428.

92. Floyd E, Rackelmann S, McQuaide S, Hartogensis W, Mehling W. Yoga for firefighters: Evaluation of a quality improvement program in California fire departments. J Bodyw Mov Ther. 2022;32:7–12. 10.1016/j.jbmt.2022.05.019.

93. Haase L, May AC, Falahpour M, Isakovic S, Simmons AN, Hickman SD, et al. A pilot study investigating changes in neural processing after mindfulness training in elite athletes. Front Behav Neurosci. 2015;9:229. 10.3389/fnbeh.2015.00229.

94. Herbert BM, Herbert C, Pollatos O, Weimer K, Enck P, Sauer H, et al. Effects of short-term food deprivation on interoceptive awareness, feelings and autonomic cardiac activity. Biol Psychol. 2012;89(1):71–9. 10.1016/j.biopsycho.2011.09.004.

95. Herbert BM, Ulbrich P, Schandry R. Interoceptive sensitivity and physical effort: implications for the self-control of physical load in everyday life. Psychophysiology. 2007;44(2):194–202. 10.1111/j.1469-8986.2007.00493.x.

96. Kosa L, Miko A, Ferentzi E, Szabolcs Z, Bogdany T, Ihasz F, et al. Body focus and cardioceptive accuracy are not associated with physical performance and perceived fatigue in a sample of individuals with regular physical activity. Psychophysiology. 2021;58(9):e13880. 10.1111/psyp.13880.

97. Koteles F, Elias I, Szabolcs Z, Kormendi J, Ferentzi E, Szemerszky R. Accuracy of reproduction of physical training load is not associated with resting heartbeat perception in healthy individuals. Biol Psychol. 2020;150:107831. 10.1016/j.biopsycho.2019.107831.

98. Machado DGDS, Farias Junior LF, Nascimento PHDD, Tavares MPM, Anselmo da Silva SK, Agricola PMD, et al. Can interoceptive accuracy influence maximal performance, physiological and perceptual responses to exercise? Physiol Behav. 2019;204:234–40. 10.1016/j.physbeh.2019.02.038.

99. Palascha A, van Kleef E, de Vet E, van Trijp HCM. The effect of a brief mindfulness intervention on perception of bodily signals of satiation and hunger. Appetite. 2021;164:105280. 10.1016/j.appet.2021.105280.

100. Parkin L, Morgan R, Rosselli A, Howard M, Sheppard A, Evans D, et al. Exploring the Relationship Between Mindfulness and Cardiac Perception. Mindfulness. 2014;5(3):298–313. 10.1007/s12671-012-0181-7.

101. Rominger C, Weber B, Aldrian A, Berger L, Schwerdtfeger AR. Short-term fasting induced changes in HRV are associated with interoceptive accuracy: Evidence from two independent within-subjects studies. Physiol Behav. 2021;241:113558. 10.1016/j.physbeh.2021.113558.

102. Schulz A, Ferreira de Sa DS, Dierolf AM, Lutz A, van Dyck Z, Vogele C, et al. Short-term food deprivation increases amplitudes of heartbeat-evoked potentials. Psychophysiology. 2015;52(5):695–703. 10.1111/psyp.12388.

103. Shen HR, Du XH, Fan YY, Dai JA, Wei GX. Interoceptive Sensibility Mediates Anxiety Changes Induced by Mindfulness-Based Tai Chi Chuan Movement Intervention. Mindfulness. 2023;14(7):1662–73. 10.1007/s12671-023-02162-z.

104. Wallman-Jones A, Palser ER, Benzing V, Schmidt M. Acute physical-activity related increases in interoceptive ability are not enhanced with simultaneous interoceptive attention. Sci Rep. 2022;12(1):15054. 10.1038/s41598-022-19235-z.

105. Kabat-Zinn J. Mindfulness-based interventions in context: Past, present, and future. Clin Psychol-Sci Pr. 2003;10(2):144–56. 10.1093/clipsy/bpg016.

106. Weng HY, Feldman JL, Leggio L, Napadow V, Park J, Price CJ. Interventions and Manipulations of Interoception. Trends Neurosci. 2021;44(1):52–62. 10.1016/j.tins.2020.09.010.

107. Britton WB. Can mindfulness be too much of a good thing? The value of a middle way. Curr Opin Psychol. 2019;28:159–65. 10.1016/j.copsyc.2018.12.011.

108. Boccia M, Piccardi L, Guariglia P. The Meditative Mind: A Comprehensive Meta-Analysis of MRI Studies. Biomed Res Int. 2015;2015:419808. 10.1155/2015/419808.

109. Cadena-Schlam L, Lopez-Guimera G. Intuitive eating: an emerging approach to eating behavior. Nutr Hosp. 2014;31(3):995–1002. 10.3305/nh.2015.31.3.7980.

110. van Strien T. Causes of Emotional Eating and Matched Treatment of Obesity. Curr Diab Rep. 2018;18(6):35. 10.1007/s11892-018-1000-x.

111. Simmons WK, DeVille DC. Interoceptive contributions to healthy eating and obesity. Curr Opin Psychol. 2017;17:106–12. 10.1016/j.copsyc.2017.07.001.

112. Davidson TL, Stevenson RJ. Appetitive interoception, the hippocampus and western-style diet. Rev Endocr Metab Disord. 2022;23(4):845–59. 10.1007/s11154-021-09698-2.

113. Lovelock DF, Tyler RE, Besheer J. Interoception and alcohol: Mechanisms, networks, and implications. Neuropharmacology. 2021;200:108807. 10.1016/j.neuropharm.2021.108807.

114. Wisniewski P, Maurage P, Jakubczyk A, Trucco EM, Suszek H, Kopera M. Alcohol use and interoception – A narrative review. Prog Neuropsychopharmacol Biol Psychiatry. 2021;111:110397. 10.1016/j.pnpbp.2021.110397.

115. Verdejo-Garcia A, Clark L, Dunn BD. The role of interoception in addiction: a critical review. Neurosci Biobehav Rev. 2012;36(8):1857–69. 10.1016/j.neubiorev.2012.05.007.

116. Baranski JV. Fatigue, sleep loss, and confidence in judgment. J Exp Psychol Appl. 2007;13(4):182–96. 10.1037/1076-898X.13.4.182.

117. Lewis KE, Blagrove M, Ebden P. Sleep deprivation and junior doctors’ performance and confidence. Postgrad Med J. 2002;78(916):85–7. 10.1136/pmj.78.916.85.

118. Forkmann T, Scherer A, Meessen J, Michal M, Schachinger H, Vogele C, et al. Making sense of what you sense: Disentangling interoceptive awareness, sensibility and accuracy. Int J Psychophysiol. 2016;109:71–80. 10.1016/j.ijpsycho.2016.09.019.

